# Method for costing a health system using a Health Systems Model

**DOI:** 10.1101/2025.01.22.25320881

**Authors:** Sakshi Mohan, Tara D. Mangal, Gerald Manthalu, Joseph Mfutso-Bengo, Margherita Molaro, Dominic Nkhoma, Bingling She, Wiktoria Tafesse, Pakwanja Desiree Twea, Simon Walker, Martin Chalkley, Tim Colbourn, Timothy B. Hallett, Andrew Phillips, Paul Revill

## Abstract

**Objectives:** Modelling approaches that consider system-wide delivery platforms rather than single diseases are increasingly recognized as crucial for the economic evaluation of policy and investment choices and can be instrumental in forward-looking policy formulation. This paper develops a costing approach tailored to one such model, the Thanzi La Onse (TLO) model of Malawi’s healthcare system, to estimate healthcare delivery costs under varying health system assumptions.

**Methods:** We developed a mixed-method costing approach to estimate the total cost of healthcare delivery in Malawi using the TLO model, from a healthcare provider perspective. Through an iterative adjustment of key costing parameters, we attempted to align our model-based estimates with real-world expenditure and budget data. Costs were estimated over an 8-year period (2023–2030) under alternative scenarios of health system capacity, including supply chain performance and the size of the health workforce.

**Results:** A detailed comparison of our cost estimates against expenditure and budget data demonstrates the reliability of our costing method and assumptions, for the conditions and resources captured by the model. Under current health system capacity, the total cost of healthcare delivery in Malawi between 2023 and 2030 was estimated at $1.53 billion [95% confidence interval, $1.51b -$1.54b], which translates to an average annual cost of $309.83 million [$306.17m -$313.56m]. The estimation of costs under alternative scenarios demonstrates the importance of capturing feedback effects to correctly forecast healthcare costs.

**Conclusion:** Mixed-method costing used within health system models, such as TLO, is a feasible and robust method for estimating healthcare delivery costs. This approach can provide valuable insights for health sector planning and resource allocation.

## 1 Introduction

There is a growing demand in global health research for modelling approaches which consider system-wide delivery platforms rather than single disease delivery platforms. These methods aim to capture the health system, its constraints and the dynamic connections between its components [1, 2, 3, 4]. All-disease health system models address this need by providing a framework to evaluate the value of diverse interventions, ranging from disease-specific to system-wide interventions, while accounting for the demographic, epidemiological, and resource interdependencies within a health system.

The use of health system models in economic evaluations requires a robust and consistent costing methodology, which consistent with the models, captures the dynamic feedback effects of changes to resource use patterns. The objective of these methods should be to estimate the cost of running the health system and implementing alternative policy choices in a way that offers a close approximation of reality.

In this paper, we develop a costing approach for the estimation of health sector costs in Malawi using the Thanzi La Onse (TLO) model, an individual-based simulation model of Malawi’s health system[5]. Through a detailed representation of resources (i.e., healthcare workers, consumables, facilities, and equipment), the population served by the public-sector health-care system, and policies that govern how resources for health care were arranged to meet healthcare needs, the TLO model provides a framework for implementing a bottom-up or micro costing method for a substantial proportion of health system inputs. The approach enables evaluations of healthcare delivery costs under alternative health system expansion paths, advancing the potential for economic evaluations to inform system-level decision-making, as well as planning for future health sector budget needs.

We present the method for estimating healthcare costs, aiming as far as possible to be representative of actual healthcare costs incurred in the country, through fidelity to local sources of unit cost information and adjustments of resource use parameters to improve the alignment of estimated costs to healthcare expenditure and budget data. We then apply this method to project total and average annual costs for the 8-year period (2023-2030) covered by the current national health sector strategy, under four scenarios representing different levels of health system capacity, specifically, the performance of consumable supply chains and the size of the health workforce.

The primary objective of this paper is to outline a method for cost estimation tailored to economic evaluations conducted using a health system model. Furthermore, by estimating resource needs in monetary terms, this approach can support with long-term budget planning and aid coordination [6], as well as guide investments in system preparedness to unforeseen health system shocks.

### Box 1

**Key concepts in the TLO model**

**Health system interaction (HSI):** This is the unit of healthcare provision, through which services are provided to patients. HSIs represent the interactions between individuals and the health system (requiring available health workers, medical consumables and equipment). These HSIs may occur at an individual’s home (in the case of community healthcare), in their neighbourhood (outreach services), or at a health facility.

**District:** One of 32 sub-national bodies (includes cities) who manage independent decentralised budgets and oversee the delivery of health within their jurisdiction [7]

**Health worker cadre:** The health workforce in the model is categorised into the following nine health worker cadres with different kinds of specialisation - clinical, nursing and midwifery, pharmacy, laboratory, dental, mental, nutrition, radiography, health surveillance assistants (HSAs)[7].

**Facility level:** Health facilities are categorised into five levels to represent their position in the referral pathway (i.e. the lowest level is that to which individuals present initially with possible onward referral to higher levels for specialised services). These levels are - health posts (level 0); health centres, maternity clinics, clinics and dispensaries (level 1a), rural or community hospitals (level 1b), District referral hospitals (level 2) and Regional referral hospitals or Central hospitals (level 3) [7].

**Disease/Public health program:** This represents a broad categorisation of individual health services into a disease or public health program which is usually managed/suprvised by independent units within the health sector [8]. For example, HIV screening, prevention and treatment services are part of the HIV program.

## 2 Methods

### 2.1 Costing methodology

#### 2.1.1 Estimation of resource use

The TLO model incorporates mechanistic representations of the causes of death and disability that, according to Global Burden of Disease (GBD) estimates, accounted for approximately 81% of deaths and 72% of disability-adjusted life-years (DALYs) in Malawi during 2015–19. The scope of the model is health care provided in Malawi during 2015–19 by the public sector, which covers 66% of all facilities and more than 98% of all outreach and village clinics in 2016. The public sector in Malawi consists of a total of 734 facilities managed by the government and the government-supported faith-based organisation (Christian Health Association of Malawi); these are grouped in the model by facility level and district. A complete description of the model can be found in Hallett et al(2024)[5].

Resource use is logged in the TLO model for every *successful* health system interaction (HSI) delivered during a simulation (see Box 1 for a description of key TLO model-related terms used in this paper). The delivery of each HSI requires an appropriate quantity of medical consumables (ie, medicines, diagnostics, and other items needed to provide care such as gloves and syringes), patient facing time (PFT) of health workers of different cadres, functional equipment, and for the facility to be in operation. Once the patient arrives at the facility to seek care, the successful delivery of an HSI depends on the availability of sufficient PFT on a simulated day, and the availability of required medical consumables and equipment. PFT is logged for every HSI which occurs before the health worker capacity for a given day at each simulated facility is exhausted.

To decide the order in which patients are seen, the ‘No Policy’ prioritisation strategy is applied, where all non-emergency cases are assigned equal priority. This means that on a given simulated day, emergency cases are attended to first (with children prioritised over adults), following which non-emergency patients are seen in a random order[9] (see Box 2 for more detail). If the HSI occurs, the quantity of consumable needed is logged when it is available; the availability of consumables is stochastically determined for each HSI based on empirical estimates of the average probability of availabilty of consumables, specific to the type of consumable, facility level, district and month of the year[10]. Similarly, the types of equipment used for each HSI are logged when they are available, based on empirical estimates from the Harmonised Health Facility Assessment 2018/19. In the current version of the TLO model (version 0.2)[11], all required equipment is assumed to always be available and functional, due to lack of granular data.

##### Box 2

**Patient Prioritisation in the TLO model** Healthcare resources are pooled in the TLO model by facility level and district, i.e. each simulated facility represents a unique combination of district and facility level (see Box 1). Within each simulated facility, constrained healthcare workers (HCWs) time is allocated using a prioritisation framework that determines the order in which patients receive care. Each patient is assigned a priority level based on the urgency of their condition. Among patients with equal priority, treatment is allocated based on how long they have been seeking care, with earlier seekers treated first. If multiple patients started seeking care on the same day, their order is randomised.

Healthcare service interactions (HSIs) are delivered sequentially each day under a rigid healthcare system assumption, meaning that services cannot be delivered if the health worker capacity for a given day runs out at a simulated facility. Patients who are not treated on the day they initially seek care can return to the same facility for up to seven days. Returning patients are treated before patients of equal priority who sought care more recently but after new patients with higher priority. [9]

#### 2.1.2 Estimation of costs

We adopt a mixed-method costing approach[12, 13], combining ingredients-based micro-costing, where feasible based on data availability, with top-down costing where necessary. Specifically, we apply micro-costing to resources that are directly attributable to service delivery and apportionable by patient. For higher-level costs, such as the distribution and warehousing of medical consumables, monitoring and supervision of health workers, and other overheads, we adopt a top-down approach. The bottom-up approach uses detailed estimates of units of each resource used for healthcare delivery. The top-down approach relies on empirical estimates of the proportion of resource costs directly used for service delivery that are allocated to such overheads within Malawi. All costs are estimated from a *healthcare provider perspective* [14], since the objective of the analysis is to guide prioritisation of investments for the Ministry of Health.

Our primary focus is to cost the *health system inputs* used for *service delivery* as represented by individual HSIs in the TLO model and any administrative expenses directly associated with these. Higher-level administrative costs (such as those for *Governance* and *Financing*) are not directly associated with service delivery and are excluded because these are outside the scope of the TLO model[3].

We cost the following *health system inputs*[15], referred to as *cost categories* - i. medical consumables, ii. human resources for health (HRH), iii. medical equipment, and iv. facility operation. We present the financial costs [4] of resources, with capital costs annuitised over the duration of use of resources such as health workers and medical equipment.

Uncertainty in our cost estimates stems from the stochastic nature of the TLO model, which simulates births, deaths, disease incidence, healthcare-seeking behaviour, and the availability of health system resources during an HSI. Following standard practice, unit costs are treated as deterministic. All costs are expressed in 2023 USD. Unit costs from previous years were appropriately inflated to 2023 values using United States Government Consumer Price Index (CPI)-based inflation rates[16]. Malawi Kwacha(MWK) values were converted to USD using exchange rates from World Development Indicators data[17].

A description of methods for estimating each cost category is provided in the sections that follow. The analysis script and input data for this study is publicly available on the TLO model repository.

##### Medical consumables

During a model simulation, every consumable *used* during an HSI is logged. This log can be aggregated at the end of the simulation to provide the total quantities of each medical consumable dispensed during each year simulated. In addition to the quantity of consumables dispensed, we also assume that some quantity is wasted due to expiration, theft or mismanagement of consumable stocks[18, 19]. We derive an empirical estimate of the proportion of consumables *wasted* in 2018 in Malawi to apply this adjustment to consumable costs. Finally, we apply a top-down estimate of supply chain costs likely to be incurred to procure, store and distribute consumables from the source to health facilities. Equations A1 - A4 explain the formulae used to calculate consumable costs.

For most consumables, we used the local prices of medical consumables in Malawi relevant to procurement by the Government of Malawi. For donated consumables, we applied market prices relevant to international donors. Details on the source of data for each component of the formula are described in table A.1.

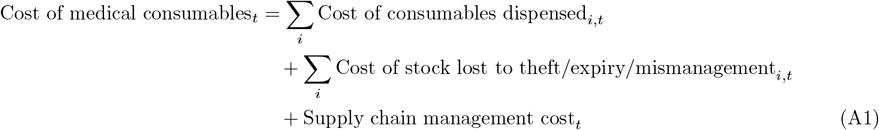

*where t = year, i = medical consumable*

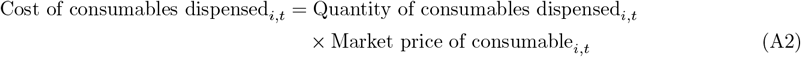

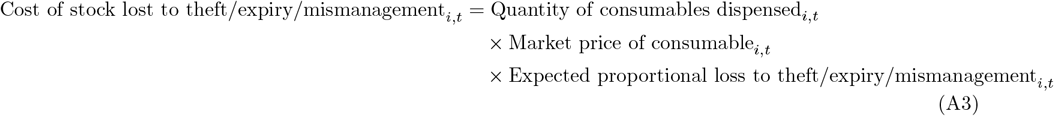

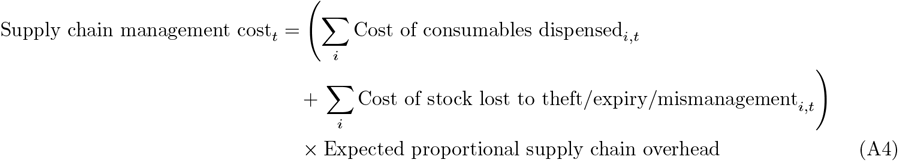

##### Human resources for Health (HRH)

Like medical consumables, the cost of employing and retaining health workers is a recurrent cost. But unlike medical consumables whose quantity purchased varies directly in proportion of the quantity of services delivered, the size of the health workforce cannot be adjusted up or down as easily through the course of the year. As such, we treat the cost of HRH as a fixed cost which can be adjusted annually based on the chosen policy of HRH expansion[4].

Following the previous HRH costing guidelines[20], our estimate of HRH costs includes remuneration, the cost of pre-service (or pre-qualification) and in-service training, and the cost of regular monitoring and supervision for all the health workers cadres included in the model. All health workers in the public sector health system are eligible for Malawi’s standard remuneration package based on staff grade. To estimate remuneration (see equation B2), we use the average annual salary of staff by cadre and facility level, weighted by the concentration of different grades within each grouping. Ideally, additional duty hours, overtime pay and pensions should also be included in this cost. However, in the absence of information on this, this is not considered in our estimates.

For pre-service training (see equation B3), we include the recurrent costs of annual tuition and salary of faculty. Following previous guidelines[20], we assume that the tuition fees would be used to cover the costs of resources (equipment and infrastructure) used in the teaching programme. we account for the salary cost of faculty to represent the opportunity cost of their time not used in delivering services. Due to a lack of data, we do not account for the costs or benefits of clinical placements of students. Similar to the approach used for costing medical equipment with multi-year durability, we annuitize the costs of training human resources over their expected working life, applying standard annuitization methodology[21]. Additionally, we account for the fact that only a proportion of students who undergo training successfully graduate, obtain a licence to practice, and are subsequently hired into the public workforce. Furthermore, some staff may be recruited from abroad^1^, bypassing the training costs within the system. For every staff member hired, a fixed recruitment cost is incurred (see equation B4).

In-service training, and monitoring and supervision costs of HR are estimated using a top-down approach as a proportion of the remuneration cost of HRH (see equations B5 and B6).

Other overheads (like management of health facilities) which enable health workers to carry out their jobs were included under *facility operation* costs.

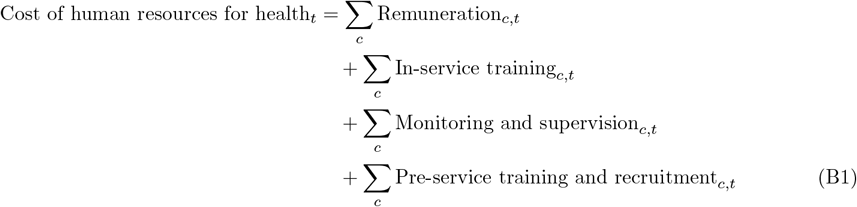

*where c = cadre, t = year*

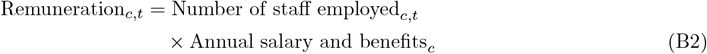

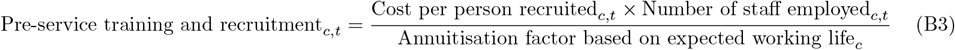

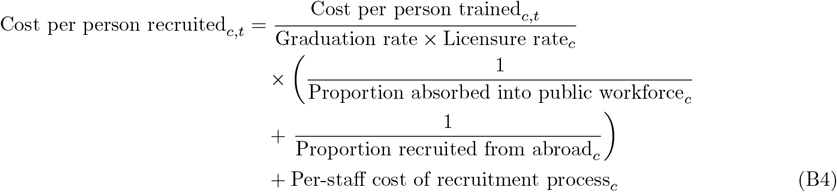

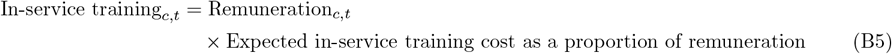

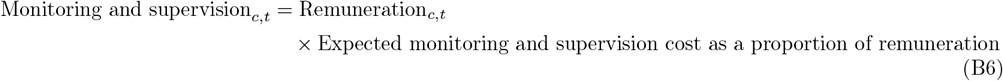

##### Medical equipment

Medical equipment is treated as a capital cost in the TLO model. Similar to medical consumables, the model logs the equipment required to deliver each HSI, and this log can be aggregated to provide a list of equipment used at each facility level during the simulation. However, unlike consumables, the model does not estimate the specific quantities of equipment required at each facility to deliver the simulated services. Instead, it only indicates whether equipment was *ever used* or *never used* at each facility level and district. To address this limitation, we rely on local expertise to determine the quantity of equipment needed by facility level. These estimates, developed to guide prioritised equipment costing in HSSP-III [22], form the basis of our calculations. Consequently, our equipment cost estimates reflect the prioritised quantities across all facilities in the country for the list of equipment identified in the simulation at each facility level. We apply the standard annuitisation method[21] to spread the cost of replacing each piece of equipment over its expected lifetime.

Crucial to ensuring the utility of medical equipment is regular servicing (including spare parts), along with ad-hoc major corrective maintenance as required[23, 24]. We use HSSP-III costing assumptions to estimate these recurrent costs as a function of the replacement cost of each equipment. See table A.1 for further information. Equations C1 - C4 describe the calculation of medical equipment costs.

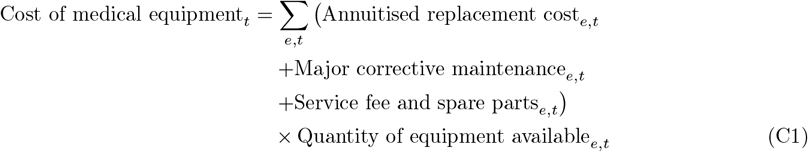

*where e = type of equipment*,

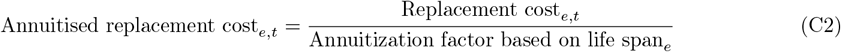

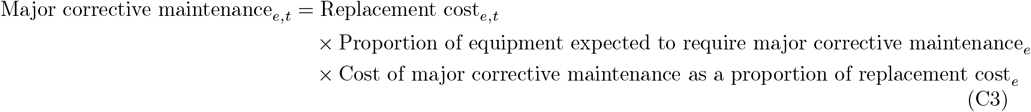

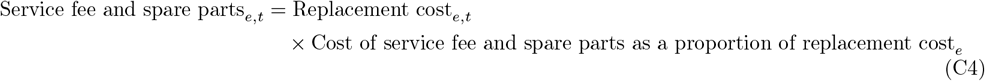

##### Facility operation costs

Recurrent costs of running health facilities are essential to service delivery and should be included in the costing exercise[15, 20]. We include the following costs in this category - utilities (electricity, water), salaries of non-healthcare delivery staff (security, cleaning, management), fuel for ambulances, overheads for inpatient services (food). These estimates are obtained from a bespoke facility audit carried out for the Thanzi La Onse model in 2024[25]. Based on data from 25 health facilities, we estimate the annual facility operation costs by facility level, as reported in table E.1.

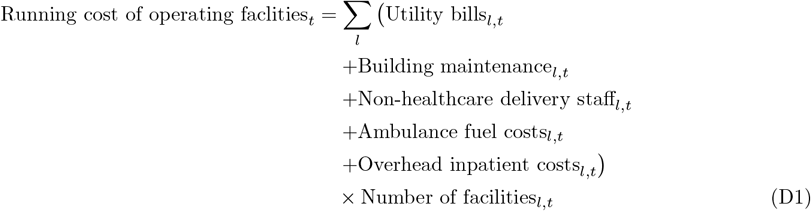

*where l = Facility level, t = year*

#### 2.1.3 Other assumptions

#### 2.1.4 Discounting

Although the standard application of a uniform 3% discount rate to future costs is now recognised as inappropriate due to variations in the projected growth rate of economies and in the projected marginal productivity of their health systems [26, 27], the majority of cost-effectiveness literature [26], including the latest WHO-CHOICE study [28, 29], continues to use the 3% discount rate. In order to align with the cost-effectiveness literature, our primary results apply the 3% annual discount rate. However, in Appendix section D, we additionally report undiscounted costs as well as costs discounted using country-specific annual projected discount rates from Lomas et al (2021)[27].

#### 2.1.5 Unit costs over time

Projecting costs over time implies that the unit prices applied to various health resources may be subject to change. For example, the cost of consumables may reduce as more generics become available, the health worker salaries may increase, the cost of fuel may fluctuate based on global price shifts, etc. In principle, the TLO model allows us to apply time-specific unit costs to relevant resource use because all interactions are located at a point in time. For the purposes of this study, however, we applied constant unit costs (as currently observed). Further, we do not account for annual inflation in our estimates and, therefore, our estimates represent *real* costs, as opposed to *nominal* costs.

For the calibration of our cost estimates with expenditure data recorded for 2018 (see section 2.1.6), we adjusted unit prices where needed to ensure that our unit costs corresponded to the unit costs relevant for that year. For example, for an annual course of antiretroviral (ARV) treatment for an adult, while the unit cost we use in the model is $37.5 [30], based on the latest prices of ARVs, for the purposes of validation, we applied a unit cost of $80 [31] relevant for 2018. The full set of unit costs can be accessed on the TLO model repository.

#### 2.1.6 Calibration of estimates

To improve confidence in our cost estimates, we compared modelled costs to available data on healthcare expenditure and budgets in Malawi. Where discrepancies were identified, we adjusted our unit cost and resource use assumptions to better align our estimates with reported data. Benchmark figures were derived using the Resource Mapping (RM) data from the latest report (see Box 3).

With the goal of bringing the model-based cost estimates within the range between reported expenditure and reported budget figures in Malawi, the following steps^2^ were followed -

1. Costs estimated from the model for the calendar year 2018 were compared to expenditure reported for the financial year (FY) July 2018–June 2019 and the maximum projected annual budget for FYs ending between 2020 and 2022^3^.
2. If the modelled cost was outside the *target range* between reported expenditure and budget, we first revisited our unit cost assumptions and extracted data from a more reliable source, if needed.
3. Next, we revisited our resource use assumptions with experts who developed the various disease modules in the TLO model. This step was only needed for medical consumables and medical equipment, because for HRH and facility operation costs, the current stock of health workers and facilities was used which was not subject to change.

- For medical consumables, where there was a significant discrepancy between RM figures and our cost estimates, we further compared the quantity dispensed logged by the model with data on actual quantity dispensed during 2018, as recorded in Open Logistics Management Information System (OpenLMIS)[32] by health facilities. If our model assumptions were found to deviate from on-the-ground practices, the number of consumable units required for each HSI was adjusted in the model to align with practice.
- For medical equipment, we compared the list of equipment logged by the model to the list of medical equipment costed in the HSSP-III [33], and added any missing equipment to the model.

The aim of this exercise was not to arbitrarily adjust parameters to match benchmark expenditures but to identify and address deviations in model assumptions from real-world conditions through the lens of costing. This approach was intended to improve the reliability and representativeness of the estimated costs. However, due to limitations in the granularity of the Resource Mapping (RM) data, we could only validate our cost estimates against categories for which health sector stakeholders reported disaggregated expenditure and budget figures. In some instances, despite the adjustments described, the modelled costs remained outside the *target range*. These discrepancies typically arose because the TLO model did not account for *all* indications that justify the use of certain resources^4^. To maintain transparency, we report these differences.

##### Box 3

**Resource Mapping (RM) Report (Round 7)** With over 60% of the health sector funding coming from external donors, and being utilised through disparate implementing partners, it can be challenging for the Ministry of Health (MOH), Malawi to keep track of resources and plan for a more efficient use of resources. To address this challenge, the MOH runs a resource mapping exercise to track expenditure and forward-looking budgets in the health sector to provide a comprehensive funding picture to help strengthen resource mobilization and resource allocation decisions for the MOH and its development partners (DPs). The RM captures data on funding for health over time, disaggregated by financing source, implementing agent, disease areas, geography, cost types and alignment to national strategic plans[34]. The round 7 data captures expenditure for the financial year(FY) ending 2019 and annual projected budgets for the FYs ending 2020, 2021, and 2022. The financial year in Malawi runs from July to June.

Although the Resource mapping report represents the best source of information on Malawi’s health sector expenditure and budgets across stakeholders, because of its self-reported nature, the quality of this data depends on the response rates and fidelity to reporting guidelines of financing and implementing partners in the sector and is, therefore, not the ultimate source of truth on the health sector financing situation[6]. In fact, it has been noted that reported expenditure tends to be far lower than the budget (due to financing challenges), and the budget better disaggregated into relavant categories than the expenditure. Therefore, we rely on the range between the two figures for the purposes of calibration, with the expenditure figure prioritised for the comparison of aggregate figures by cost category.

### 2.2 Projection of costs under different health system assumptions

To demonstrate how the cost of service delivery changes interactively through changes in health system capacity/performance, we use the costing method described above to project healthcare resource use costs under four scenarios, varying the performance of consumable supply chains and size of the health workforce. We make these projections for the 8-year period covered by the HSSP-III (2023–2030).

1. **Actual:** This is the baseline scenario which aims to represent the *actual* capacity of the health system in Malawi as observed between 2018 and 2022. This includes - i. current probability of availability for each *medical consumable* at each facility level and district[32], ii. current stock of *human resources for health (HRH)* [7], iii. current *performance of healthcare workers* in clinical diagnoses and probability of referrals[5], and iv. current propensity for *healthcare seeking* [35]. The *default* probability of availability of consumables of 49.2% on average (44% at level 1a and 57% at level 1b facilities). Details on the size of health workforce can be found in She et al (2024)[7].
2. **Expanded HRH:** Ceteris paribus, this scenario expands the number of health workers at the rate of 6% annually starting in 2020.
3. **Improved consumable availability:** Ceteris paribus, this scenario increases the probability of consumable availability in health facilities at levels 1a and 1b (see Box 1), where most of the health care is delivered, to match that of the 75th percentile of top-performing facilities [10], at the corresponding facility level for each individual consumable^5^. This results in an 8 percentage points and 10 percentage points increase in the average probability of availability of consumables at levels 1a and 1b respectively, in comparison to the *actual* scenario. See Figures B.1 and B.2 for a detail on the improvement in consumable availability under this scenario by *program*.
4. **Expanded HRH + Improved consumable availability:** This scenario combines the health system improvements from scenarios 2 and 3.

## 3 Results

### 3.1 Performance of costing model: comparison of 2018 cost estimates to actual expenditure and budget

The total health sector expenditure reported in Malawi for FY ending 2019 was $624.1 million. Excluding components which are outside the scope of the TLO model[3], i.e. programme management and administration, newly built infrastructure, infrastructure upgrades, purchase of motor vehicles, and medical consumables for conditions not included in the Essential Health Package (EHP)[36] of Malawi, the expenditure was $404 million. Based on the TLO model, we estimate the total healthcare cost to be $358.9 million (11.2% lower than the RM expenditure estimate).

Zooming in, figure 1 provides a snapshot of the TLO model cost estimate plotted against annual expenditure and budget estimates in Malawi for each of the four cost categories. We find that for most of the RM categorisations, our cost estimates are within the *target range* between the actual expenditure and budget figures. Appendix C provides a detailed breakdown of this comparison and discusses the potential source of discrepancies.

**Figure 1:**
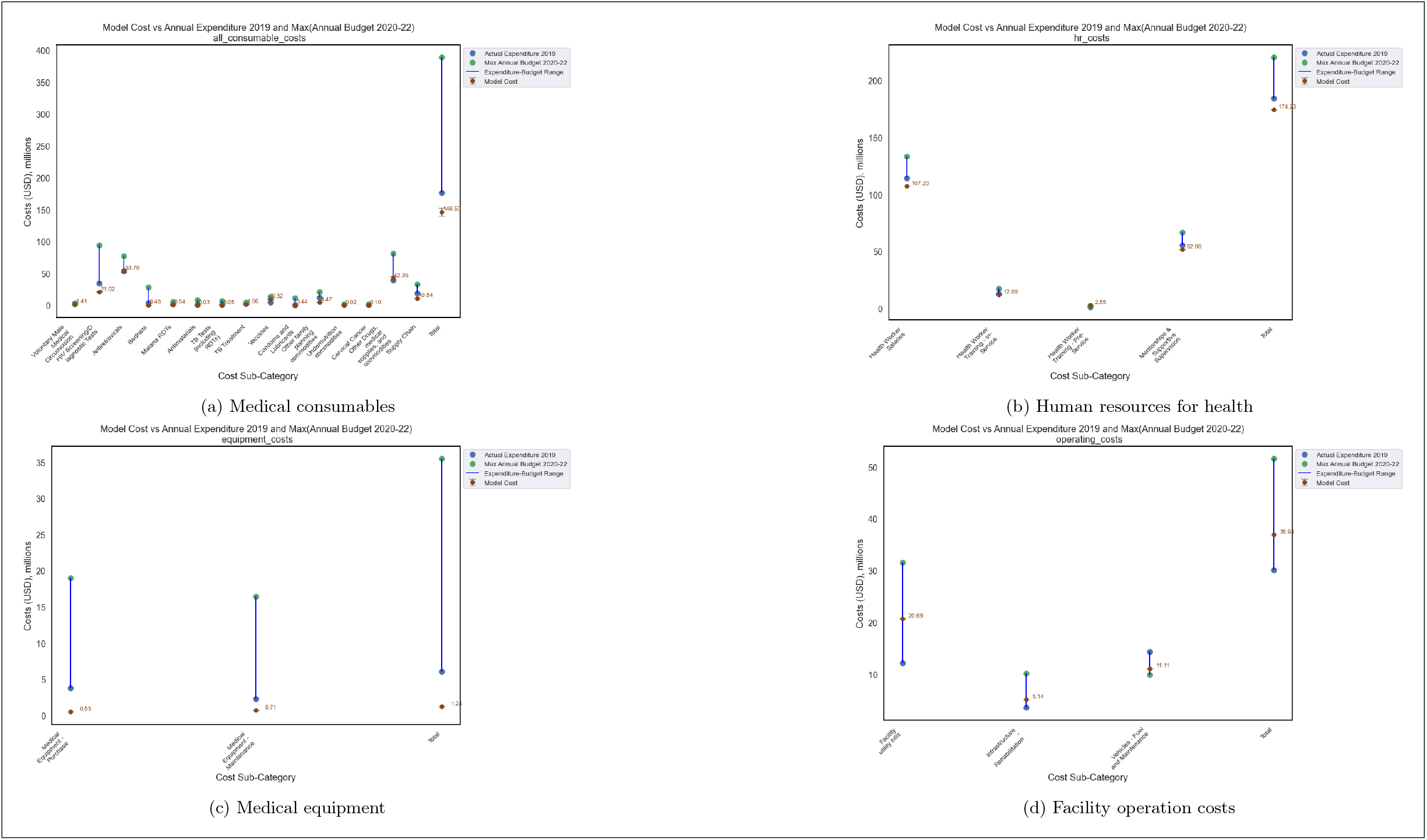
Comparison of model-based cost estimates with actual expenditure recorded for 2018/19 and budget planned for 2019/20-2021/22: For each cost-category, we compare our cost estimates by a comparable disaggregated category as per Malawi’s Resource Mapping data. The goal of this exercise was for our estimated costs to be with the range between annual expenditure and annual budget, if justifiable adjustments to resource use and unit costs could be made to improve the alignment of our estimates to data.

### 3.2 Projection of costs under different health system assumptions, 2023-2030

The total cost of healthcare delivery in Malawi between 2023 and 2030 was estimated to be $1.53 billion [95% confidence interval (CI), $1.51b - $1.54b], under the actual scenario, and increased to $1.54 billion [$1.52b - $1.56b] under the improved consumable availability scenario, followed by $2.10 billion[$2.08b - $2.12b] under the expanded HRH scenario and finally $2.44 billion[$2.43b - $2.46b] under the expanded HRH + improved consumable availability scenario. This translates to an average annual cost of $309.83 million[$306.17b - $313.56b], under the actual scenario, $313.08 million [$309.23b - $316.96b] under the improved consumable availability scenario, followed by $429.40 million [$425.03b - $433.56b] under the expanded HRH scenario and finally $499.01 million [$495.89b - $502.15b] under the expanded HRH + improved consumable availability scenario. Figure 2 breaks these estimates down by cost category and figure 3 reports these by subcategory. Figure 4 reports these costs by year between 2023 and 2030.

**Figure 2:**
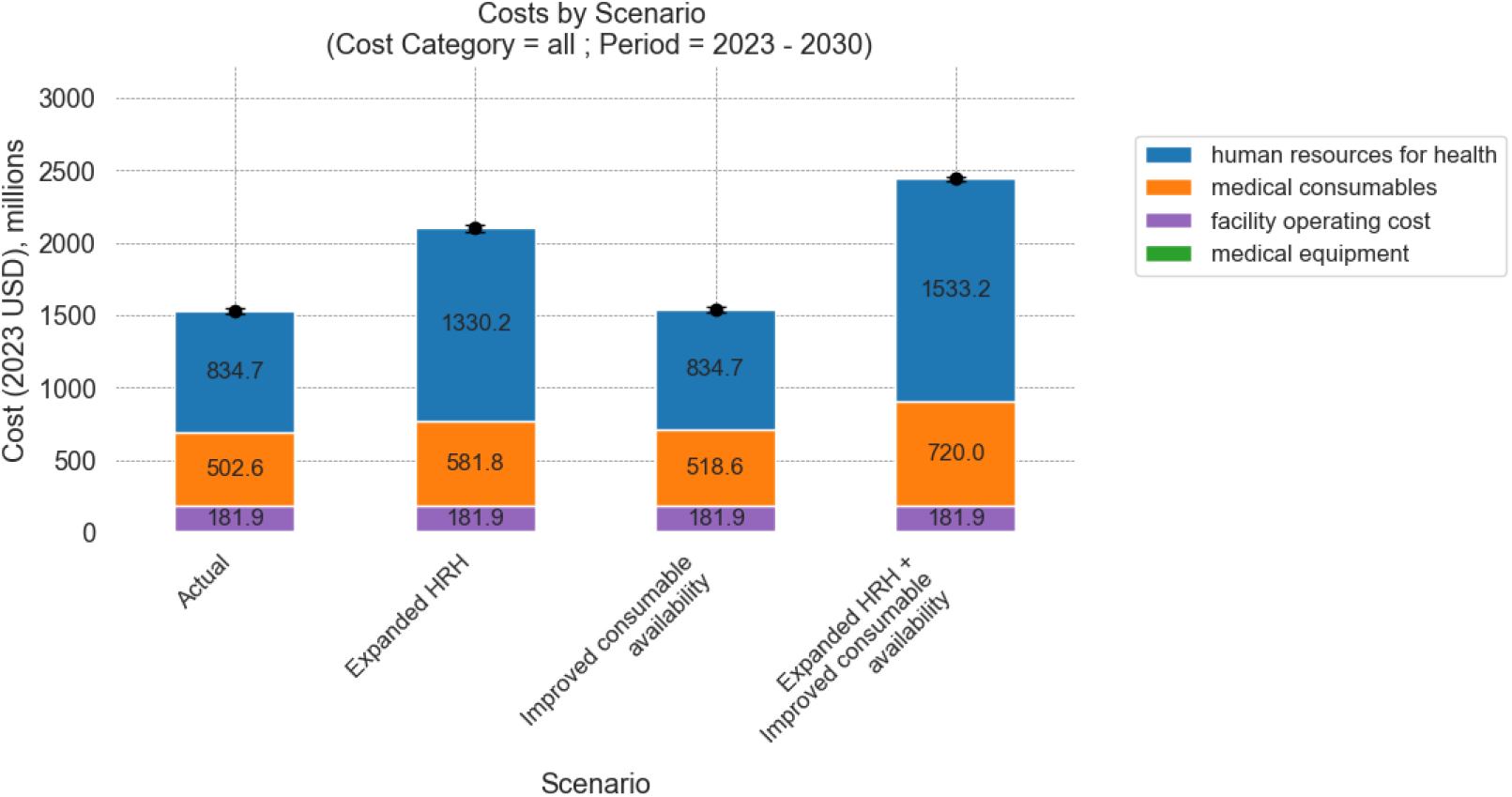
Estimated costs by cost category, 2023-2030. This figure presents the total cost by cost category for 8 years under four scenarios modelled. The black error bars represent the 95% confidence interval around the total cost. The values are measured in 2023 USD and discounted at 3% to obtain the NPV in 2023. Note that uncertainty in our cost estimates stems from the stochastic nature of demographic, epidemiological and resource availability parameters in the TLO model; unit costs are assumed to be deterministic.

**Figure 3:**
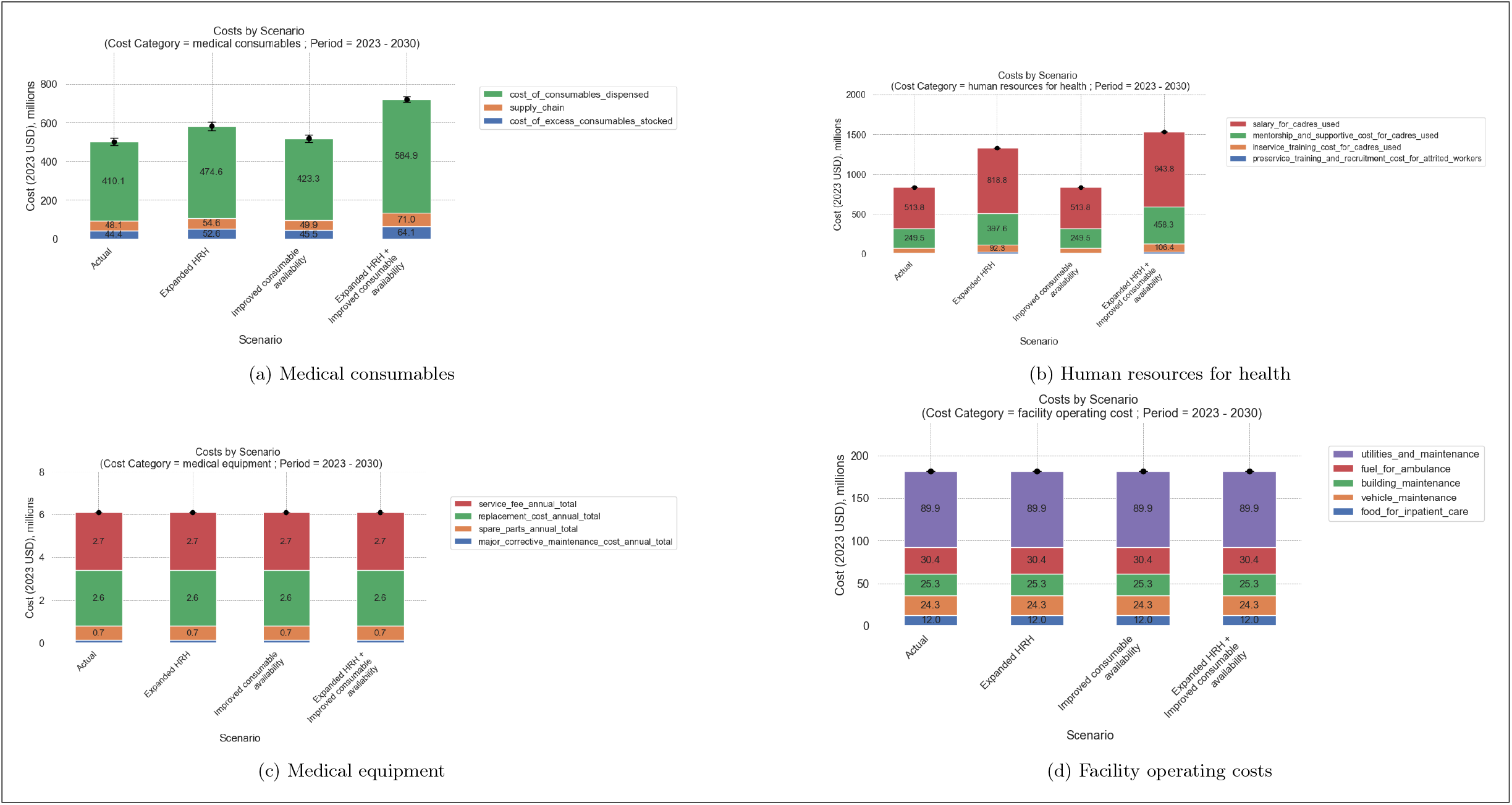
Estimated cost by category and subcategory, 2023-2030. This figure presents the total cost by category and subcategory for 8 years under four scenarios modelled. The values are measured in 2023 USD and discounted at 3%.

**Figure 4:**
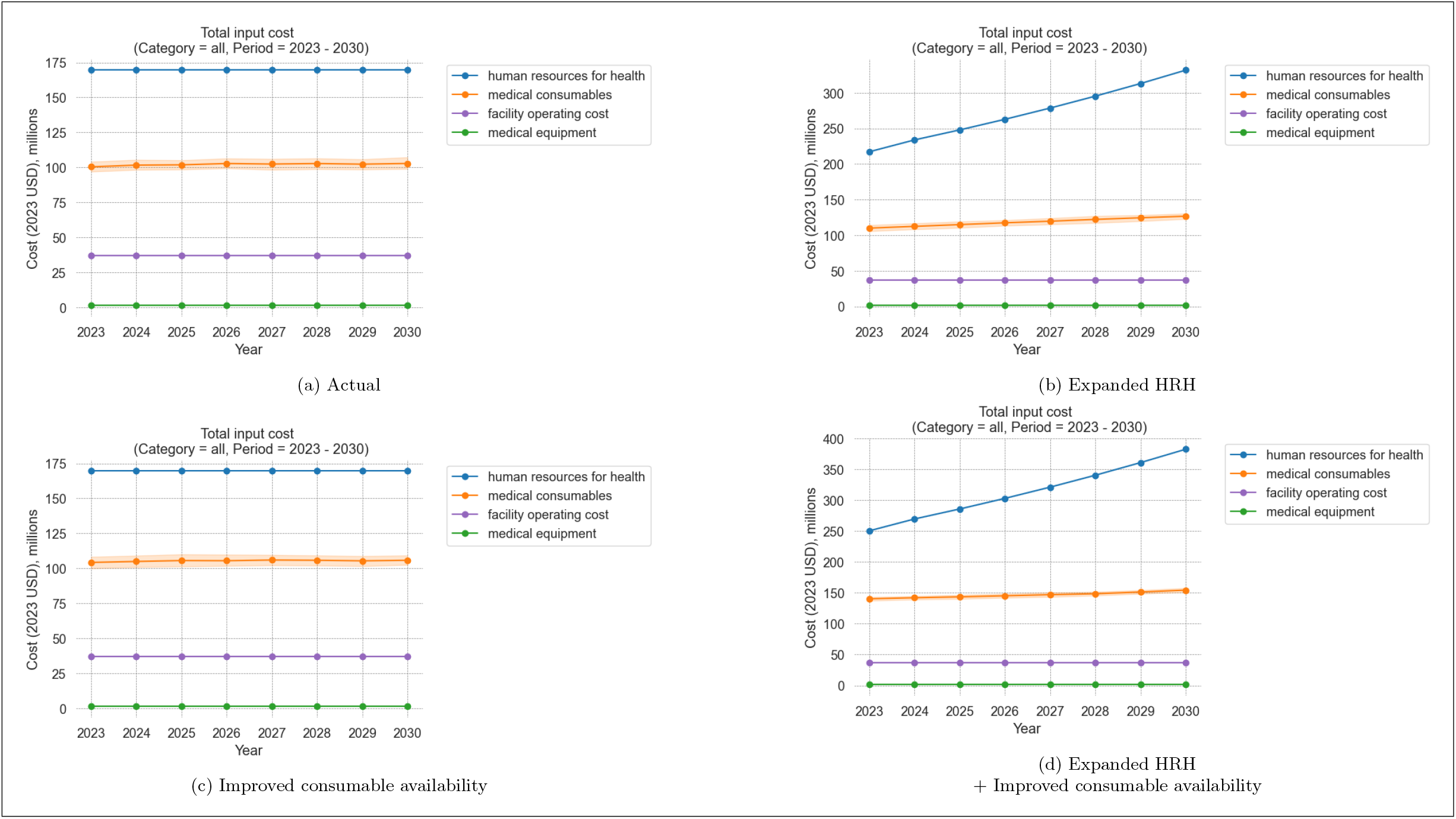
Estimated cost by year, 2023-2030. This figure presents the total cost by category for 8 years under four scenarios modelled. The values are measured in 2023 USD and undiscounted values of cost are reported for each year.

Notably, we find that the *improved consumable availability* scenario results in a 15.77% increase in cost of medical consumables. However, when combined with expanded HRH, the increase in consumables cost is 43.26% more than the *actual* scenario because the health system is able to deliver more appointments.

A detailed breakdown of costs by subcategory and category-specific subgroups (e.g., consumables for consumable costs, health worker cadre for HR costs) is available in Appendix F. Figures F.1 - F.4 summarize these breakdowns for the top 10 subgroups ranked by cost. These figures illustrate how the allocation towards the top 10 medical consumables as a percentage of total cost shifts across different scenarios, reflecting variations in the types of health services successfully delivered under different health system capacities. For instance, the cost of HIV testing consumables increases by 7.96% under the expanded HRH scenario and by 323.9% under the combined expanded HRH and improved consumable availability scenario, while showing almost no change under the scenario with improved consumable availability alone, highlighting the interdependence between human resources and consumable availability in delivering high-impact interventions like HIV testing. In contrast, the distribution of costs across subgroups for human resources for health (HRH), medical equipment, and facility operations remains constant under all scenarios, suggesting that these cost categories are less sensitive to the system-level changes considered in the scenarios modelled in this study.

These estimates use a 3% annual discount rate. Undiscounted cost estimates and those with variable discount rates from Lomas et al (2021)[27] are reported in appendix D.

## 4 Discussion

In this analysis, we used the Thanzi La Onse (TLO) model to estimate current and future costs of healthcare delivery in Malawi under different health system capacity assumptions. We demonstrated how mixed-method costing can be applied to a mechanistic health system model and where appropriate data can be found to cost various health system inputs.

While calibration to epidemiological data is a well-established practice for individual-based models[37], there are no standardized methods for calibrating cost and resource use estimates to data[38]. Much of the previous work projecting healthcare costs into the future has relied on top-down costing approaches, rather than ingredients-based approaches centred on resource use [39, 40, 41, 42]. Consequently, there is limited guidance on target benchmarks for the calibration of cost estimates, particularly when using bottom-up costing methods. We make an important contribution in this space by demonstrating how an iterative unit cost and resource-use adjustment process can be used to align resource use assumptions and model-based cost estimates to practice and real-world data.

We were able to align our cost estimates with Malawi’s resource mapping data for most cost components through -i. the use of country-specific (or donor-specific when relevant) unit cost data for the costing for health resources, ii. calibration of epidemiological, demographic and health system productivity parameters to actual observed data, and iii. an iterative process of revisiting TLO model assumptions on resource use parameters. Any deviations from the data were reported to enhance the use and interpretation of our results, and potential causes of these deviations were discussed.

Using our representative costing method, we project the cost of healthcare delivery in Malawi under different assumptions about health system capacity. The TLO model uniquely captures the feedback effects of expanding one health system component on others, a key strength of our approach. These findings highlight the importance of understanding the interdependence of health system components when forecasting future costs. This costing method provides a solid foundation for evaluating policy decisions in Malawi, particularly those involving interventions with dynamic feedback effects.

We present a flexible costing framework capable of accommodating greater complexity. However, as applied in this paper, the approach has the following limitations:

1. For some cost components, we used a top-down approach, assuming constant returns to scale and scope, which may not hold as the system expands significantly in the future. For instance, we assumed supply chain costs are a fixed proportion of medical consumable purchases. However, interventions focused on strengthening the supply chain would require more detailed costing, including labor, storage, procurement, and distribution costs of consumables [43]. In the future, appropriate non-linear scaling functions may also be applied to such costs[44].
2. By representing the availability of consumables as a probability parameter for each facility on a given day and costing only the consumables dispensed when available, the cost of consumables in the *improved health system* scenario is higher than in the *actual* scenario. However, this approach does not account for the fact that ensuring timely consumable availability depends not only on the total quantity purchased but also on distribution efficiency. Our method does not capture variations in availability due to differences in supply chain efficiency.
3. At this stage, the TLO model does not account for the actual availability of equipment at health facilities. Instead, we adopted a normative approach, assuming that the equipment requirements outlined by the HSSP-III are met at all facilities. In reality, just as a health worker can only see a fixed number of cases per day, medical equipment like an X-ray machine can only handle a limited number of cases daily. Ideally, our costing method would estimate the minimum quantity of equipment needed at each facility based on its maximum utilization rate (or throughput). However, estimating maximum throughput is challenging, and future studies could reference tools like the WHO Essential Inputs Estimator tool, which provides estimates for equipment needs per bed or facility size [45]. Additionally, while we apply standard assumptions for recurring service fees and equipment maintenance, our method does not reflect the actual functionality of equipment in the country, which is expected to be below normative standards, based on regional evidence [24].
4. In the improved health system scenarios, we only capture the cost of recruiting/purchasing additional resources. Any overheads incurred to *implement* such a change, along with any accompanying health system investments (such as additional facility space) that would be needed to make the resource expansion effective, are excluded from our estimates. In other words, *implementation costs* of step changes to the health system are not included in our analysis.
5. We assume no variation in unit cost by facility ownership (government or faith-based organisations) and region.
6. Given the large number of unit cost parameters involved and the limited evidence on their uncertainty, we adopted a deterministic approach for applying unit costs to our estimates. In reality, unit costs may fluctuate across consumables, equipment, facility levels, districts, and years.

Despite these limitations, our analysis underscores the robustness and adaptability of our costing approach for the TLO model. By integrating diverse data sources and using a comprehensive mixed-methods approach, we provide a valuable framework for assessing the financial implications of various policy decisions in Malawi’s health sector. This framework can support evidence-based prioritization, financial planning and advocacy, and resource allocation. As such, this work forms part of a significant step forward in leveraging health systems modelling to inform policy and strengthen health sector planning.

## Data Availability

All data produced in the present work are contained in the manuscript.

https://github.com/UCL/TLOmodel/blob/sakshi/costing/resources/costing/ResourceFile_Costing.xlsx

## Appendix

### A. Data Sources

**Table A.1:** Sources of costing data.

| Parameter | Data Source |
| --- | --- |
| **Medical consumables** |  |
| Quantity of consumables dispensed$_{i,t}$ | A estimate of the number of units dispensed of each medical consumable included in the TLO model is generated by a model simulation. This estimate is based on a detailed log of the quantity of consumable 'used' during each health system interaction. Note that a consumable is logged as *used* only when it is available at a health facility; any unmet *requests*, due to stock-outs for consumables are not logged. |
| Market price of consumable$_{i,t}$ | The main reference for the price of consumables is the 2023 drug price catalogue of the Central Medical Stores Trust (CMST)[46]. Established by the Government of Malawi in 1968, CMST is the institution responsible for procuring and distributing medical consumables to health facilities. Since 2011, it has been operating as a fully autonomous not-for-profit trust[47]. When the price of a drug was not found in the CMST catalogue, we referred to the Essential Healthcare Package Tool (2021) used to guide service prioritisation for the Health Sector Strategic Plan (HSSP)-III of Malawi[48], or the One Health Tool (2016) used to guide the development of the HSSP-II. The price of consumables usually purchased by donors from international suppliers is obtained from relevant references based on the program - HIV/AIDS consumables[30], TB consumables[49], nutrition and family planning consumables[50].<br>Certain medical supplies have unique characteristics which do not lend themselves well to the standard cost estimation method. These *separately-managed supplies* are either produced or collected within the health system, or are delivered through a platform independent of health facilities. In this subcategory, we include the cost of oxygen, blood, and indoor residual spraying (IRS) for malaria prevention. Estimates of the per unit cost of these supplies are obtained from literature, including administration costs. The cost of oxygen is assumed to be \$0.00057 per litre based on Malawi's One Health Tool (2016)[51]. For blood, we use the estimate of \$155 per unit from Mafirakureva et al (2016)[52], which includes the cost of recruitment, collection, testing, processing, storage and distribution of blood. This estimate is from Zimbabwe and is used as a proxy for the likely costs in Malawi. For IRS, we apply a cost of \$2.58 per person protected, including the cost of personnel, equipment, fuel, logistics and planning, shipping and PPE, from a study based in Tanzania[53]. |

**Table A.1:** Summarized Costs by Category and Category Subgroup.

| Parameter | Data Source |
| --- | --- |
| Expected proportional loss to theft/expiry/mismanagement$_{i,t}$ | Estimates of drug wastage based on literature range from 14% to 35%[54, 18]. Notably, there is considerable variation in wastage across programs [18]. To obtain granular estimates of the extent of wastage for each medical consumable, we used data from Malawi’s Open Logistics Management Information Systems (OpenLMIS) records from 2018. The OpenLMIS is a monthly record maintained by all government and government-supported faith-based health facilities in Malawi of the quantity of consumables in stock, received and dispensed along with the number of days of stock-out experienced during the month. Using this data, we are able to estimate the total *inflow* of each consumables into health facilities during 2018 (excluding what remained in stock at the end of 2018 and was transferred to the stock for 2019) and the total *outflow*. The ratio of the inflow to outflow minus 1 gives us the extent of wastage of each consumable. The average wastage was estimated to be 14.3%, ranging between 2% for insecticide-treated nets to 70% for bleomycin powder. See figure E.1 for a summary of inflow to outflow ratios by disease/public health program category of consumables.<br>Similar to other consumables, we assume that some quantity of *separately managed supplies* may be wasted. In the absence of data on wastage specific to these consumables, we applied the average rate of wastage across all consumables (14%) to calculate the total cost of these supplies. |
| Expected proportional supply chain overhead | Based on data on health sector expenditure from the Health Sector Resource Mapping Report Round 7 (2022)[55], we assume that the supply chain overhead costs (warehousing, storage and transportation) are 11.23% of the purchase cost of consumables. This is comparable to previous estimates from literature[56]. |
| **Human resources for Health** |  |
| Annual salary and benefits$_c$ | We estimate annual salary and benefits for each of the nine health worker cadres by level of care. The salary of a staff member belonging to a specific cadre is determined by their grade. Using the detailed healthcare staff returns data from Malawi, we calculated the average annual salary by cadre and level of care, weighted by the number of staff employed at each grade for each cadre and level. |
| Number of staff employed$_{c,t}$ | This is obtained from the HSSP-III [22]. For more detail, refer to She et al (2024)[7]. When an gradual or instantaneous increase in the size of the workforce is modelled, the number of health workers costed in each year may vary. |
| Expected in-service training cost as a proportion of remuneration$_c$ | Based on data on health sector expenditure from the Health Sector Resource Mapping Report Round 7 (2022) [55], we assume that the cost of in-service training is 11.27% of the annual remuneration cost for every cadre. |
| Expected monitoring and supervision cost as a proportion of remuneration$_c$ | Based on data on health sector expenditure from the Health Sector Resource Mapping Report Round 7 (2022)[55], we assume that the cost of in-service training is 48.55% of the annual remuneration cost for every cadre. |
| Expected working life$_c$ | Based on the expected annual attrition rate of 7% assumed in the costing exercise for the HSSP-III[22], the expected working life of a health worker in the public health sector is assumed to be 14.3 years. Although we would expect this to vary regionally and by cadre, due to lack of data, we assume the same expected working life across all health workers. |
| Cost per person trained$_{c,t}$ | We estimate the cost per person trained as the cost of annual tuition (assuming these are subsidised by the government) plus the annual remuneration cost of the faculty required to run the training program times the average duration of the program. Using data from the HSSP-III HRH Costing tool[22], we estimate the former for every cadre by taking the average tuition fee across medical training institutes in Malawi, weighted by their enrollment rate. To estimate the remuneration cost of faculty, we use the ratio of students to faculty for each relevant faculty cadre from HSSP-III assumptions to estimate the faculty remuneration cost per student. |

Table A.1: Summarized Costs by Category and Category Subgroup
| Parameter | Data Source |
| --- | --- |
| Graduation rate<sub>c</sub> | Assumed to be 80% following HSSP-III costing assumptions [22]. |
| Licensure rate<sub>c</sub> | Assumed to be 90% following HSSP-III costing assumptions [22]. |
| Proportion absorbed into public workforce<sub>c</sub> | Based on HSSP-III costing assumptions[22], this varies from 61% for the pharmacist cadre to 100% for community health workers. |
| Proportion recruited from abroad<sub>c</sub> | Assumed to be 0% following HSSP-III costing assumptions [22]. |
| Per-staff cost of recruitment process<sub>c</sub> | Assumed to be \$33.75 per staff recruited following HSSP-III costing assumptions [22]. |
| **Medical Equipment** |  |
| Replacement cost<sub>e</sub> | Estimates of the replacement cost of equipment were extracted directly from HSSP-III costing tools[33]. |
| Life span<sub>e</sub> | Life span of each type of equipment was extracted from the UK government’s Department for International Development (DFID)-commissioned ‘How to Plan and Budget for Your Healthcare Technology’ report[23] |
| Proportion of equipment expected to require major corrective maintenance<sub>e</sub> | Assumed to be 20% for all equipment over 8 years following HSSP-III costing assumptions [22]. |
| Cost of major corrective maintenance as a proportion of replacement cost<sub>e</sub> | Assumed to be 20% of the cost of the equipment requiring major corrective maintenance, following HSSP-III costing assumptions [22]. |
| Cost of service fee/spare parts as a proportion of replacement cost<sub>e</sub> | Service fee assumed to be 10% of the value of the equipment annually and spare parts assumed to cost 2.5% of the value of the equipment annually, following HSSP-III costing assumptions [22]. |
| Quantity of equipment available<sub>e</sub> | The HSSP-III set out an aspirational and a prioritised quantity of each type of equipment which are required at each facility level. Due to lack of access to data on the actual equipment inventory, we used the prioritised inventory size to cost medical equipment. |
| Number of facilities<sub>l</sub> | The TLO model represents a total of 734 government and Christian Health Association of Malawi (CHAM) facilities in Malawi grouped by facility level and district [5]. For the purpose of costing equipment we multiply the equipment cost per facility by the number of facilities at each facility level. |
| **Facility operation costs** |  |
| Estimates of annual operation costs (monthly minimum, average, and maximum during 2024) were collected through a facility audit survey under the Thanzi La Mawa project. The estimates were aggregated by facility level. See table E.2. In line with other deterministic unit costs used, we only used the mean value of the average monthly costs reported in our cost estimation. |  |

### B. Status of availability of consumables under *Actual* and *Improved consumable availability* scenarios

**Figure B.1:**
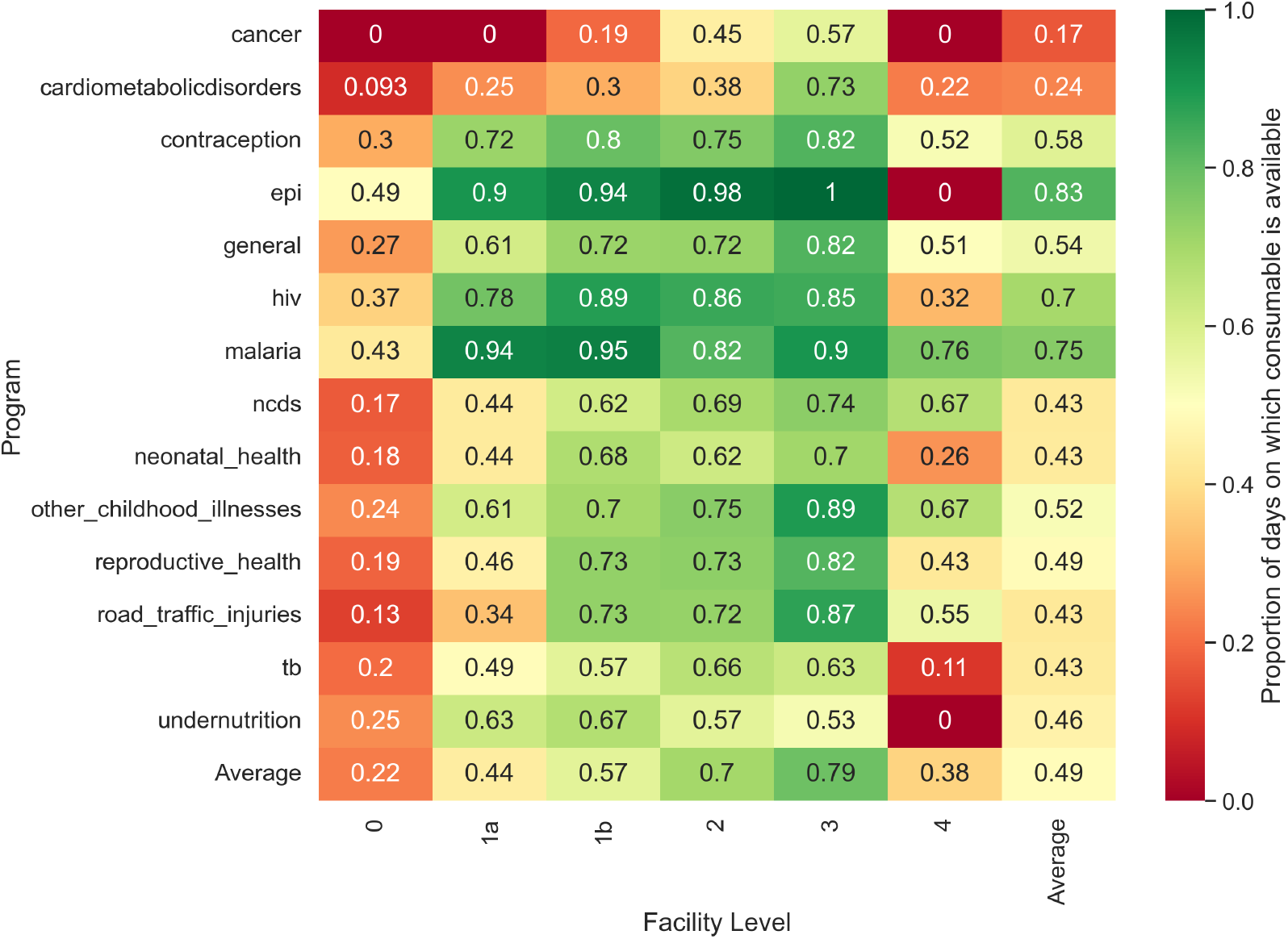
Average probability of consumable availability by program, *actual* scenario.

**Figure B.2:**
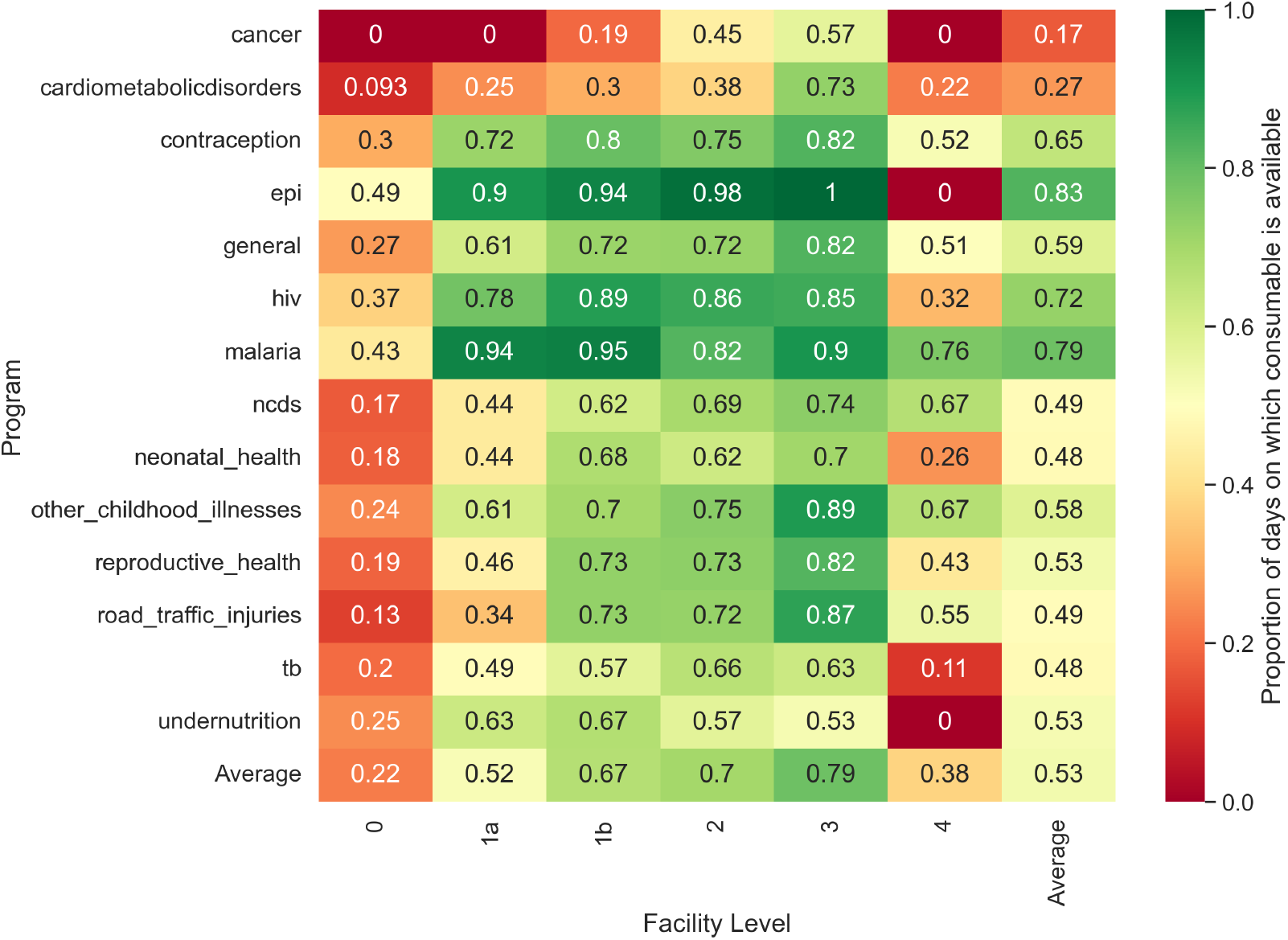
Average probability of consumable availability by program, *improved consumable availability* scenario.

### C. Comparison of Model Estimates with Resource Mapping data

**Table C.2:** Comparison of Model Estimates with Resource Mapping data.

| Cost Category | Relevant RM group | Recorded Expenditure (FY 2018/19) | Maximum Recorded Annual Budget (FY 2019/20 - 2021/22) | Estimated cost (TLO Model, 2018) | Deviation of estimated cost from actual expenditure (%) |
| --- | --- | --- | --- | --- | --- |
| Not represented in TLO model | Infrastructure - New Builds | 29,663,284.53 | 6,768,215.83 | NA | NA |
| Not represented in TLO model | Infrastructure - Upgrades | 8,782,296.75 | 8,832,821.37 | NA | NA |
| Not represented in TLO model | Non-EHP consumables | 25,178,739.81 | 18,706,715.51 | NA | NA |
| Not represented in TLO model | Program Management & Administration | 137,153,224.50 | 140,499,397.41 | NA | NA |
| Not represented in TLO model | Supply Chain - non-EHP consumables | 2,598,793.35 | 4,587,061.15 | NA | NA |
| Not represented in TLO model | Unclassified | 0.00 | 3,622,078.57 | NA | NA |

Table C.2: Comparison of Model Estimates with Resource Mapping data
| Cost Category | Relevant RM group | Recorded Expenditure (FY 2018/19) | Maximum Recorded Annual Budget (FY 2019/20 - 2021/22) | Estimated cost (TLO Model, 2018) | Deviation of estimated cost from actual expenditure (%) |
| --- | --- | --- | --- | --- | --- |
| Not represented in TLO model | Vehicles - Fuel and Maintenance (Beyond Governm... | 8,309,105.90 | 21,629,947.92 | NA | NA |
| Not represented in TLO model | Vehicles - Purchase | 8,392,198.49 | 5,145,880.06 | NA | NA |
| facility operating cost | Facility utility bills | 12,142,707.98 | 31,595,108.55 | 20,692,033.51 | Within target range |
| facility operating cost | Infrastructure - Rehabilitation | 3,586,443.58 | 10,177,394.16 | 5,137,488.70 | Within target range |
| facility operating cost | Vehicles - Fuel and Maintenance | 14,370,090.62 | 9,865,240.53 | 11,111,862.64 | Within target range |
| human resources for health | Health Worker Salaries | 114,418,952.79 | 133,228,884.25 | 107,230,088.91 | -6.28% |
| human resources for health | Health Worker Training - In-Service | 12,897,136.55 | 17,743,710.12 | 12,086,818.36 | -6.28% |
| human resources for health | Health Worker Training - Pre-Service | 1,277,697.67 | 2,665,199.15 | 2,852,164.56 | 123.23% |
| human resources for health | Mentorships & Supportive Supervision | 55,553,332.80 | 66,652,165.02 | 52,062,955.21 | -6.28% |
| medical consumables | Antimalarials | 681,848.59 | 8,094,056.65 | 25,609.00 | -96.24% |
| medical consumables | Antiretrovirals | 53,796,360.12 | 77,506,903.49 | 53,787,113.67 | -0.02% |
| medical consumables | Bednets | 3,552,600.00 | 28,061,308.64 | 463,707.63 | -86.95% |
| medical consumables | Cervical Cancer | 534,872.32 | 1,581,977.74 | 101,750.65 | -80.98% |
| medical consumables | Condoms and Lubricants | 210,313.84 | 11,211,211.72 | 442,117.95 | Within target range |
| medical consumables | HIV Screening/Diagnostic Tests | 34,469,044.13 | 94,345,358.80 | 21,024,664.01 | -39.00% |
| medical consumables | Indoor Residual Spray | 7,272,001.18 | 13,194,497.82 | NA | NA |
| medical consumables | Malaria RDTs | 1,719,000.00 | 5,643,435.92 | 538,367.86 | -68.68% |
| medical consumables | Other Drugs, medical supplies, and commodities | 39,437,013.14 | 80,801,692.06 | 42,346,013.41 | Within target range |
| medical consumables | Other family planning commodities | 12,323,143.36 | 21,052,691.19 | 4,468,052.84 | -63.74% |
| medical consumables | Supply Chain | 18,742,481.24 | 33,081,856.02 | 10,835,411.02 | -42.19% |
| medical consumables | TB Tests (including RDTs) | 599,011.62 | 6,695,563.77 | 53,719.77 | -91.03% |
| medical consumables | TB Treatment | 2,338,536.34 | 4,685,956.00 | 1,664,427.22 | -28.83% |
| medical consumables | Undernutrition commodities | 913,214.58 | 1,721,063.60 | 23,268.04 | -97.45% |
| medical consumables | Vaccines | 4,200,418.52 | 13,209,399.57 | 9,319,174.55 | Within target range |
| medical consumables | Voluntary Male Medical Circumcision | 2,891,681.00 | 1,832,645.01 | 1,408,451.89 | -51.29% |
| medical equipment | Medical Equipment - Maintenance | 2,294,331.38 | 16,468,941.94 | 712,772.18 | -68.93% |
| medical equipment | Medical Equipment - Purchase | 3,754,149.89 | 19,024,579.29 | 525,327.86 | -86.01% |

**Note:** This is still work in progress and some consumable and medical equipment costs are being adjusted.

### D Costs under different discount rates Figure

**Figure D.1:**
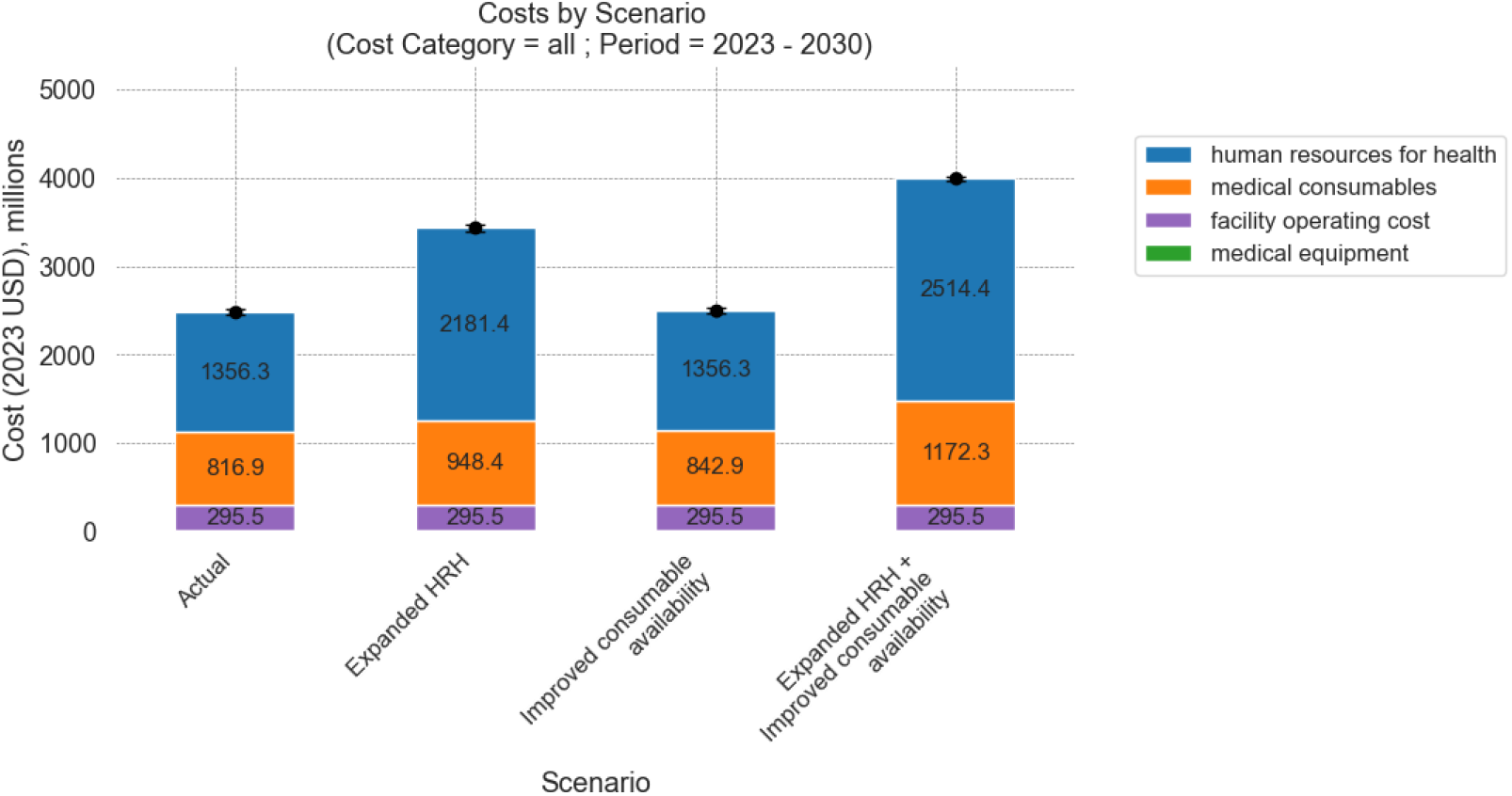
Estimated costs by cost category (undiscounted), 2023-2030. This figure presents the total cost by category for 8 years under four scenarios modelled. The black error bars represent the 95% confidence interval around the total cost. The values are measured in 2023 USD and are aggregated over the period without discounting.

**Figure D.2:**
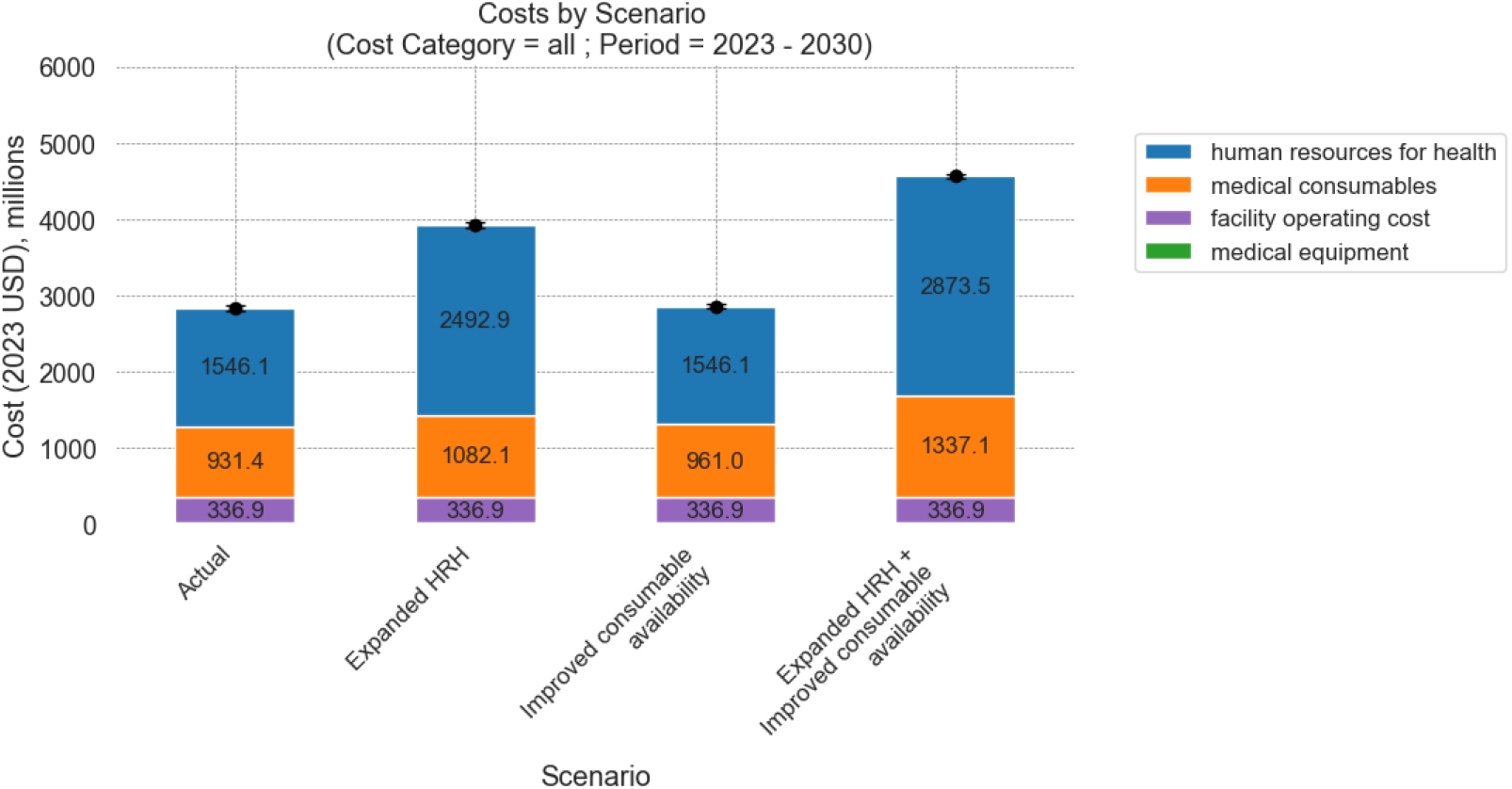
Estimated cost by cost category (Lomas et al (2021) discount rates), 2023-2030: This figure presents the total cost by category for 8 years under four scenarios modelled. The values are measured in 2023 USD and are aggregated over the period based on Malawi-specific discount rates changing over time[27]. Note that after accounting for the low projected GDP growth rate and an expected positive trend in the marginal productivity of the health system (*k*), the discount rate for health (*r*_*h*_) for Malawi was projected to be negative. Hence, these values are larger than the undiscounted costs reported in Figure D.1.

### E Selected cost input data

**Figure E.1:**
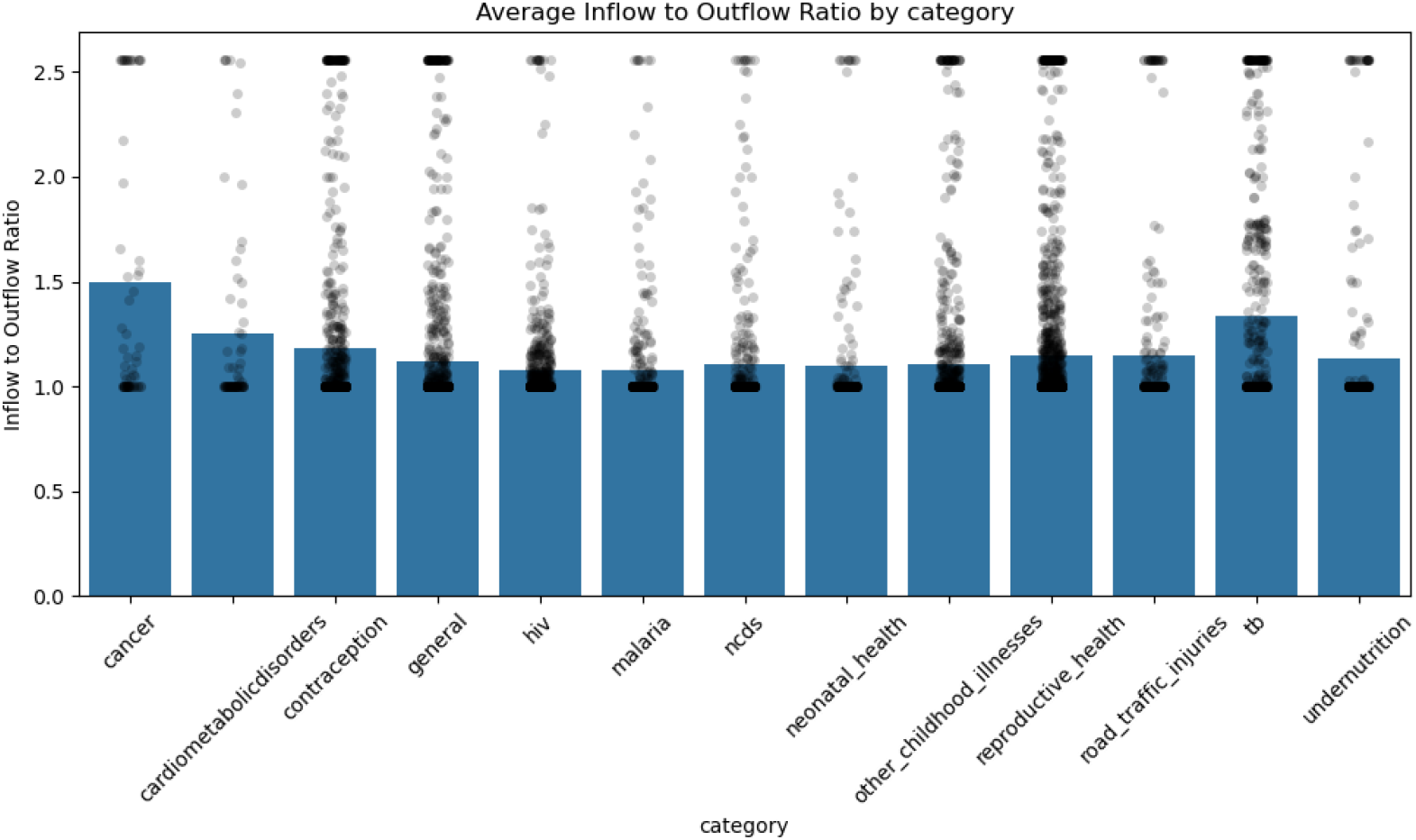
Ratio of medical consumable inflow to outflow, 2018: This figure shows the average rate of wastage of consumables by disease/health program, based on monthly facility reports on consumables received and dispensed. The black dots represent the inflow to outflow ratio by consumable, facility level, and district. The top 5% values have been trimmed and equated to the 95th percentile value. The bars represent the average ratio of inflow to outflows across districts and facility levels within a category.

**Table E.1:** Annual unit cost used for cost estimation of facility operations. These represent 12 times the mean of the average monthly cost for each type of operating cost reported by facilities in the TLM survey. See table E.2 for a detailed summary of the data collected.

| Facility Level | Electricity | Water | Cleaning | Security | Building maintenance | Facility management | Vehicle maintenance | Ambulance fuel | Food for inpatient cases |
| --- | --- | --- | --- | --- | --- | --- | --- | --- | --- |
| 1a | 12,215.86 | 1,828.93 | 1,163.55 | 1,838.70 | 5,654.00 | 1,195.16 | 5,085.15 | 4,171.55 | - |
| 1b | 11,691.69 | 11,436.70 | 9,308.83 | 21,061.16 | 21,116.53 | 8,278.09 | 24,541.47 | 42,668.24 | 18,999.06 |
| 2 | 51,713.44 | 30,775.24 | 17,375.72 | 16,046.16 | 9,285.44 | 16,720.68 | 18,961.59 | 39,819.35 | 32,866.76 |
| 3 | 174,417.48 | 141,679.72 | 65,866.60 | 52,512.42 | 163,178.26 | - | 51,755.33 | 96,163.90 | 159,441.04 |

**Table E.2:**
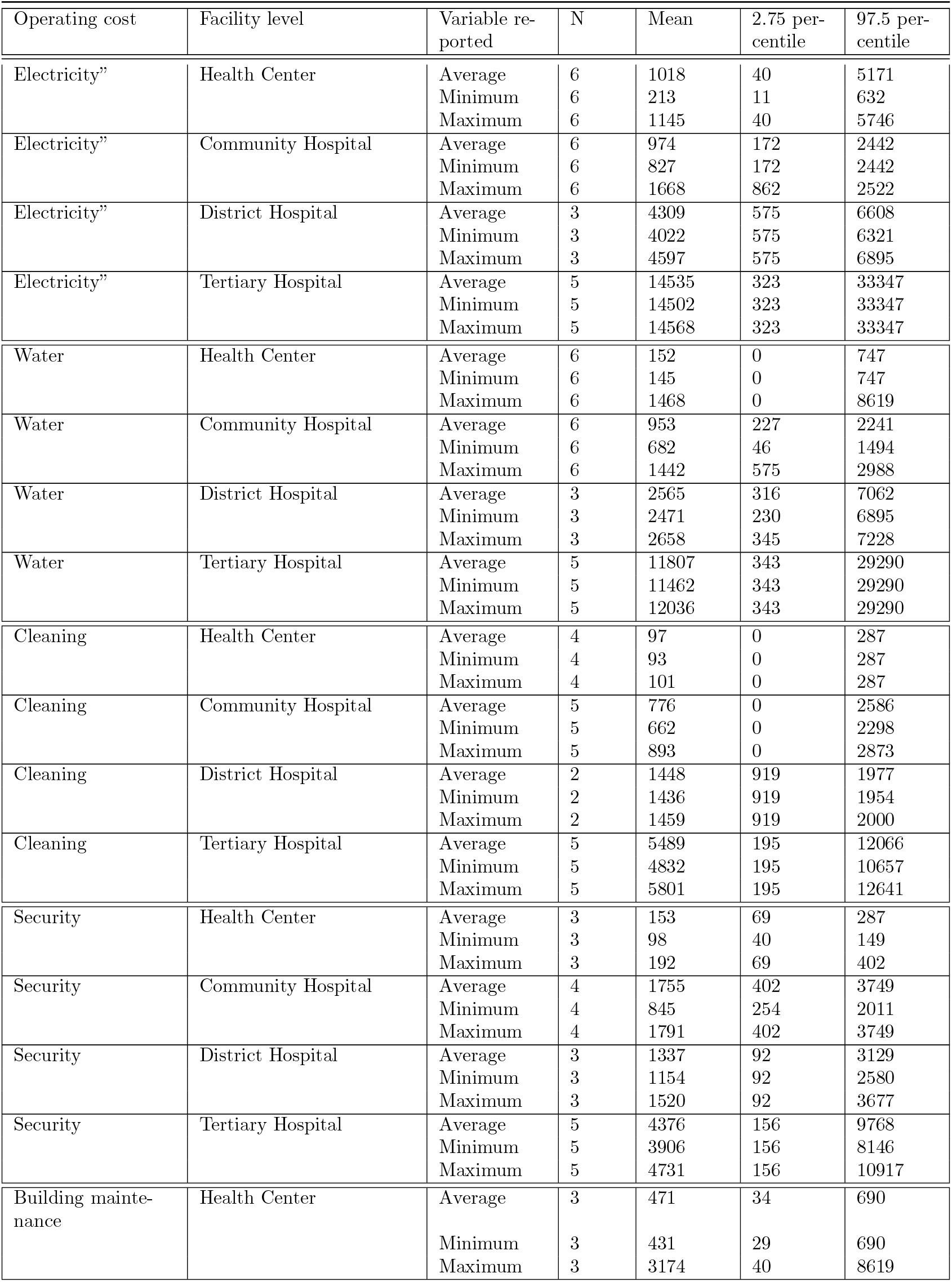
Summary of facility operating costs reported in the TLM Datasets.

**Table E.2:** Detailed data on facility operating costs reported in the TLM Datasets.

| Operating cost | Facility level | Variable reported | N | Mean | 2.5 percentile | 97.5 percentile |
| --- | --- | --- | --- | --- | --- | --- |
| Building maintenance | Community Hospital | Average | 6 | 1760 | 105 | 6119 |
|  |  | Minimum | 6 | 1090 | 3 | 5056 |
|  |  | Maximum | 6 | 3011 | 105 | 7182 |
| Building maintenance | District Hospital | Average | 3 | 774 | 23 | 1867 |
|  |  | Minimum | 3 | 678 | 23 | 1724 |
|  |  | Maximum | 3 | 870 | 23 | 2011 |
| Building maintenance | Tertiary Hospital | Average | 5 | 13598 | 97 | 37349 |
|  |  | Minimum | 5 | 10569 | 97 | 28730 |
|  |  | Maximum | 5 | 15478 | 97 | 40222 |
| Facility management | Health Center | Average | 3 | 100 | 0 | 184 |
|  |  | Minimum | 3 | 100 | 0 | 184 |
|  |  | Maximum | 3 | 100 | 0 | 184 |
| Facility management | Community Hospital | Average | 4 | 690 | 0 | 2183 |
|  |  | Minimum | 4 | 661 | 0 | 2069 |
|  |  | Maximum | 4 | 719 | 0 | 2298 |
| Facility management | District Hospital | Average | 2 | 1393 | 488 | 2298 |
|  |  | Minimum | 2 | 1106 | 488 | 1724 |
|  |  | Maximum | 2 | 1681 | 488 | 2873 |
| Facility management | Tertiary Hospital | Average | 1 | 0 | 0 | 0 |
|  |  | Minimum | 1 | 0 | 0 | 0 |
|  |  | Maximum | 1 | 0 | 0 | 0 |
| Vehicle maintenance | Health Center | Average | 4 | 424 | 0 | 776 |
|  |  | Minimum | 4 | 395 | 0 | 776 |
|  |  | Maximum | 4 | 2485 | 0 | 8619 |
| Vehicle maintenance | Community Hospital | Average | 4 | 2045 | 115 | 7470 |
|  |  | Minimum | 4 | 1727 | 102 | 6321 |
|  |  | Maximum | 4 | 2440 | 259 | 8619 |
| Vehicle maintenance | District Hospital | Average | 3 | 1580 | 718 | 2298 |
|  |  | Minimum | 3 | 1436 | 575 | 2298 |
|  |  | Maximum | 3 | 1724 | 862 | 2298 |
| Vehicle maintenance | Tertiary Hospital | Average | 4 | 4313 | 1403 | 8379 |
|  |  | Minimum | 4 | 3663 | 140 | 8379 |
|  |  | Maximum | 4 | 4528 | 1403 | 8379 |
| Ambulance Fuel | Health Center | Average | 4 | 348 | 0 | 977 |
|  |  | Minimum | 4 | 244 | 0 | 575 |
|  |  | Maximum | 4 | 402 | 0 | 1149 |
| Ambulance Fuel | Community Hospital | Average | 5 | 3556 | 116 | 13216 |
|  |  | Minimum | 5 | 3019 | 116 | 12641 |
|  |  | Maximum | 5 | 4091 | 116 | 14365 |
| Ambulance Fuel | District Hospital | Average | 3 | 3318 | 603 | 8547 |
|  |  | Minimum | 3 | 3266 | 517 | 8475 |
|  |  | Maximum | 3 | 3371 | 690 | 8619 |

| Operating cost | Facility level | Variable reported | N | Mean | 2.5 percentile | 97.5 percentile |
| --- | --- | --- | --- | --- | --- | --- |
| Ambulance Fuel | Tertiary Hospital | Average | 5 | 8014 | 276 | 20111 |
|  |  | Minimum | 5 | 7369 | 276 | 17238 |
|  |  | Maximum | 5 | 8646 | 276 | 22984 |
| Food for inpatient care | Health Center | Average | 3 | 0 | 0 | 0 |
|  |  | Minimum | 3 | 0 | 0 | 0 |
|  |  | Maximum | 3 | 0 | 0 | 0 |
| Food for inpatient care | Community Hospital | Average | 3 | 1583 | 613 | 2528 |
|  |  | Minimum | 3 | 1195 | 195 | 1781 |
|  |  | Maximum | 3 | 1938 | 930 | 3275 |
| Food for inpatient care | District Hospital | Average | 3 | 2739 | 402 | 4654 |
|  |  | Minimum | 3 | 2586 | 402 | 4482 |
|  |  | Maximum | 3 | 2892 | 402 | 4827 |
| Food for inpatient care | Tertiary Hospital | Average | 5 | 13287 | 495 | 30209 |
|  |  | Minimum | 5 | 12931 | 495 | 30209 |
|  |  | Maximum | 5 | 13758 | 495 | 30209 |

### F Detailed cost outputs

#### F.1 Cost by category-specific subgroup

**Table F.1:**
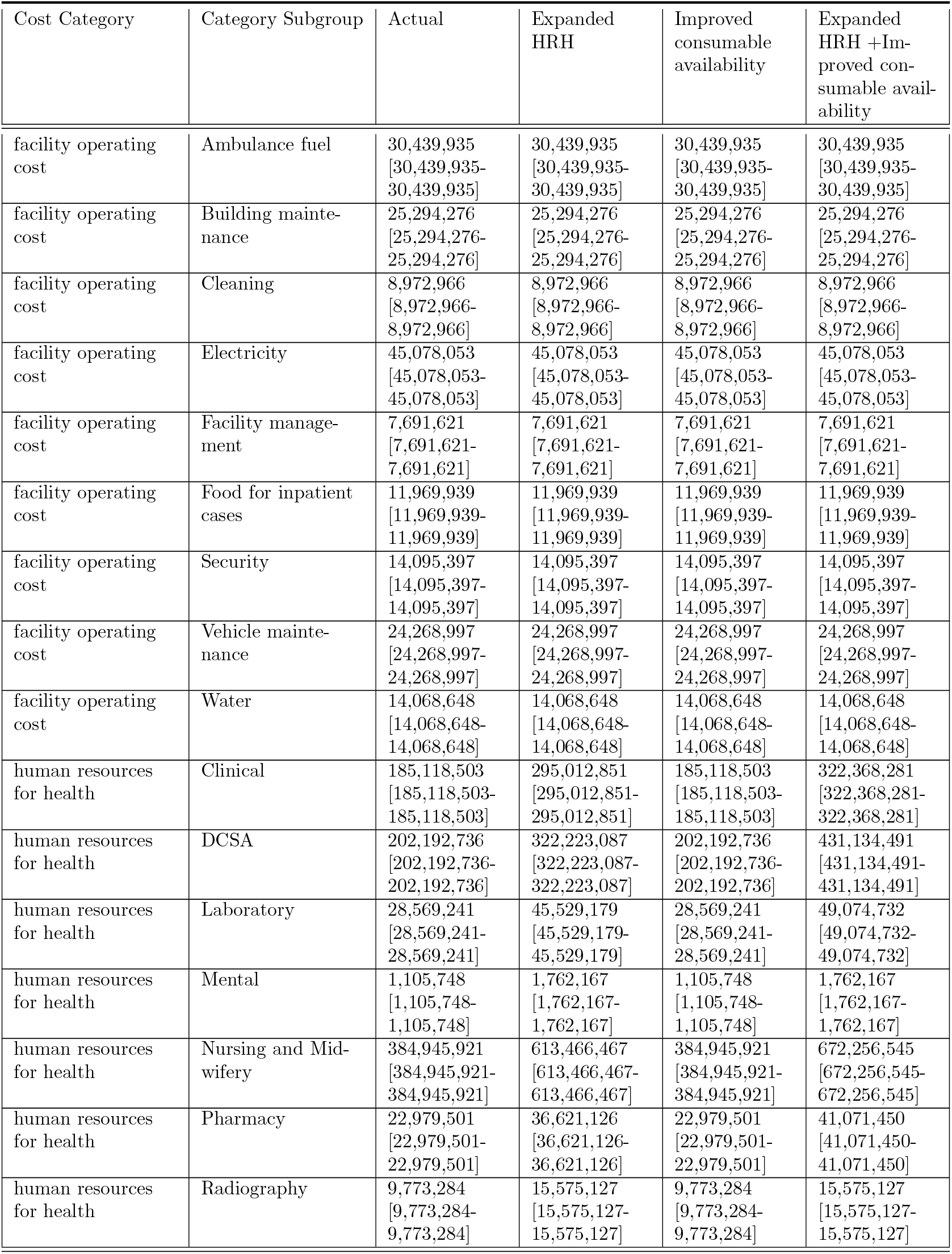
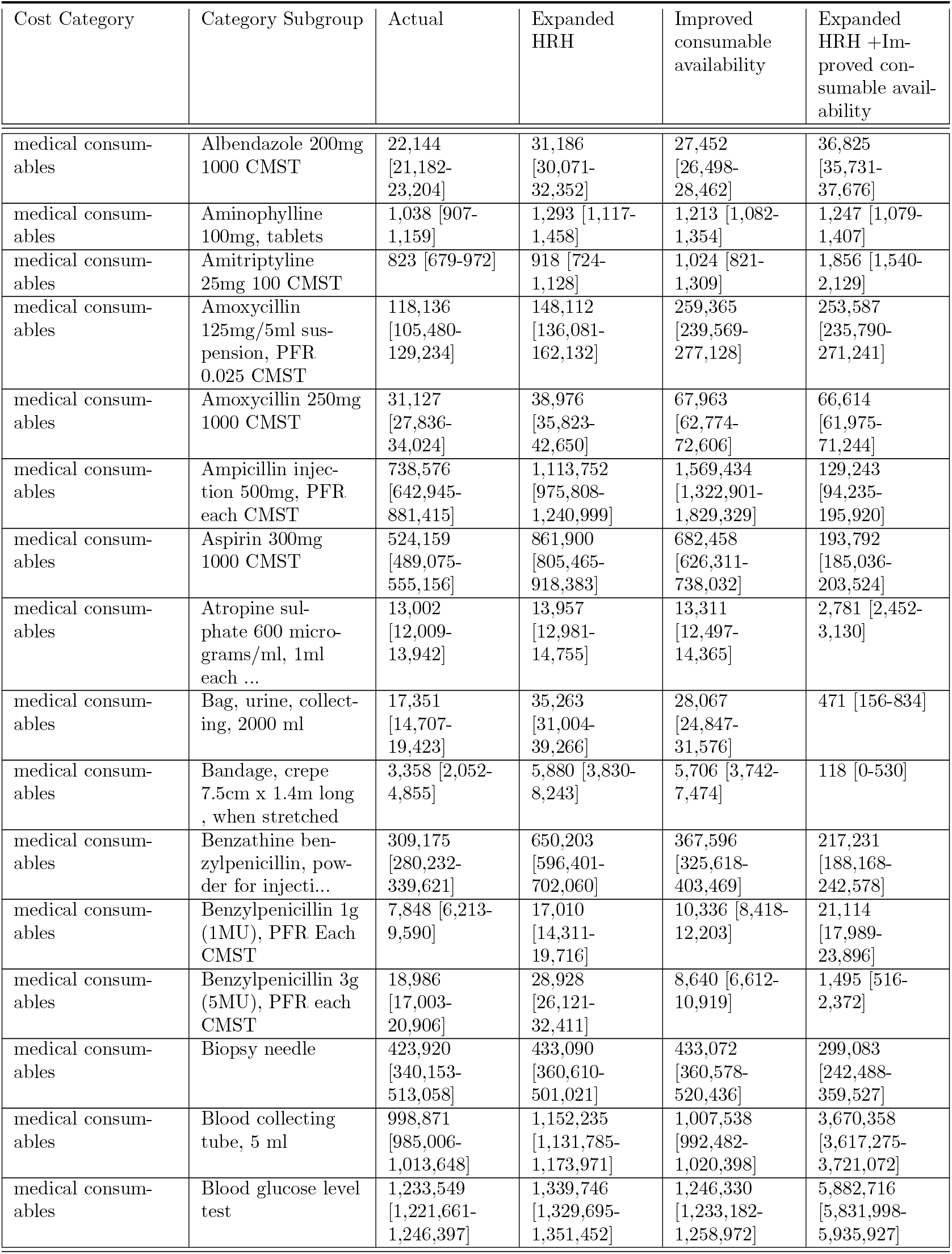
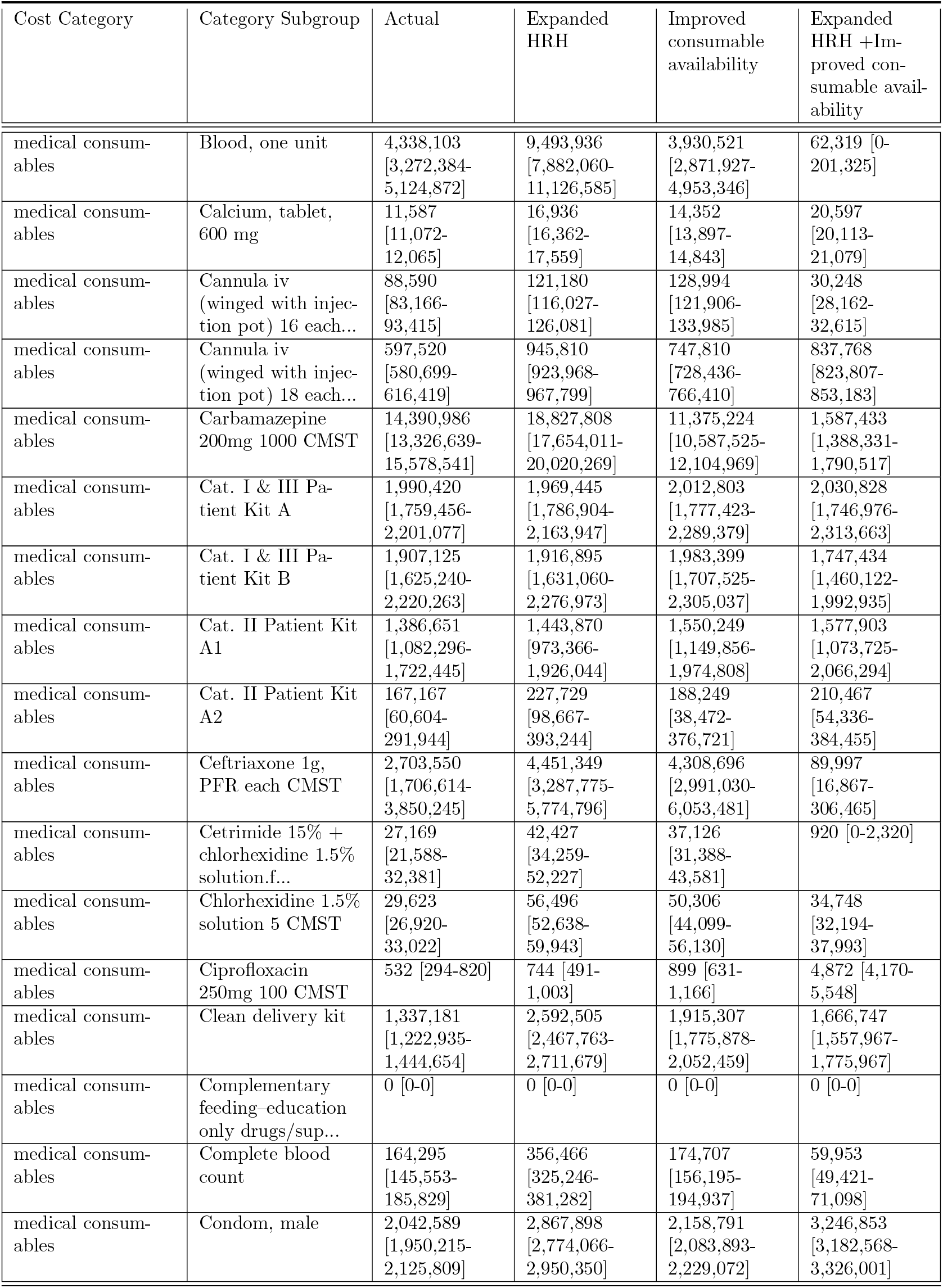
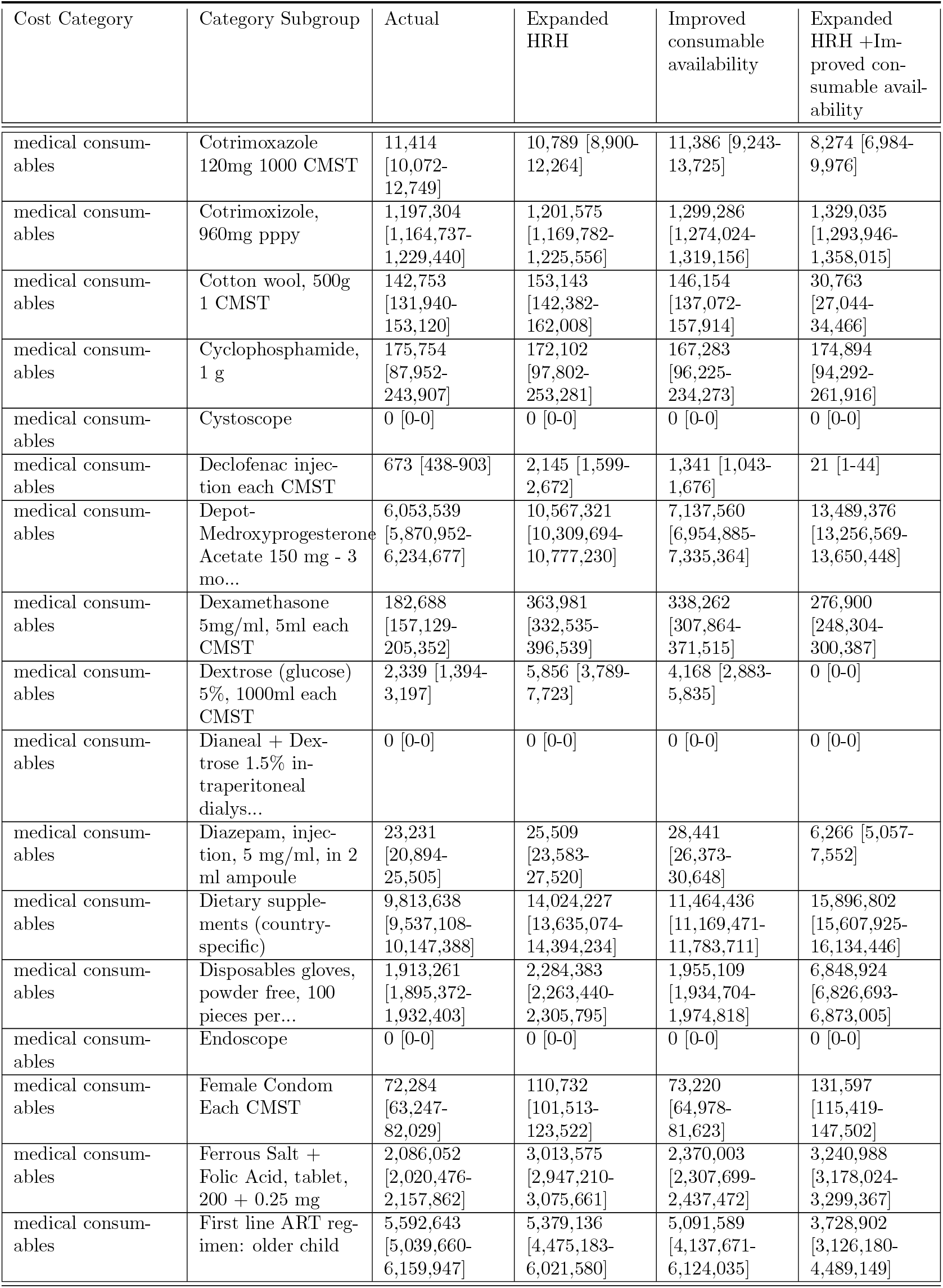
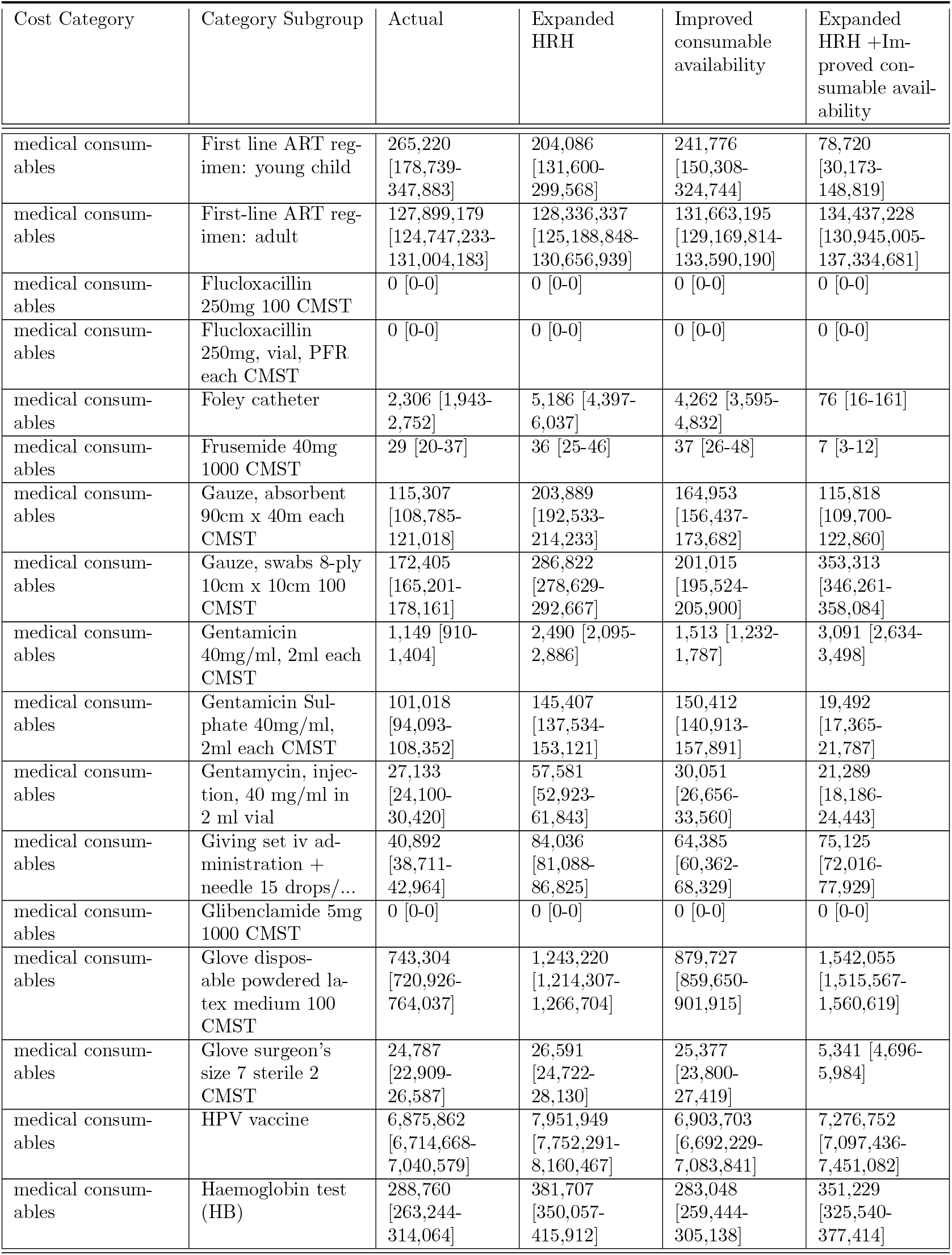
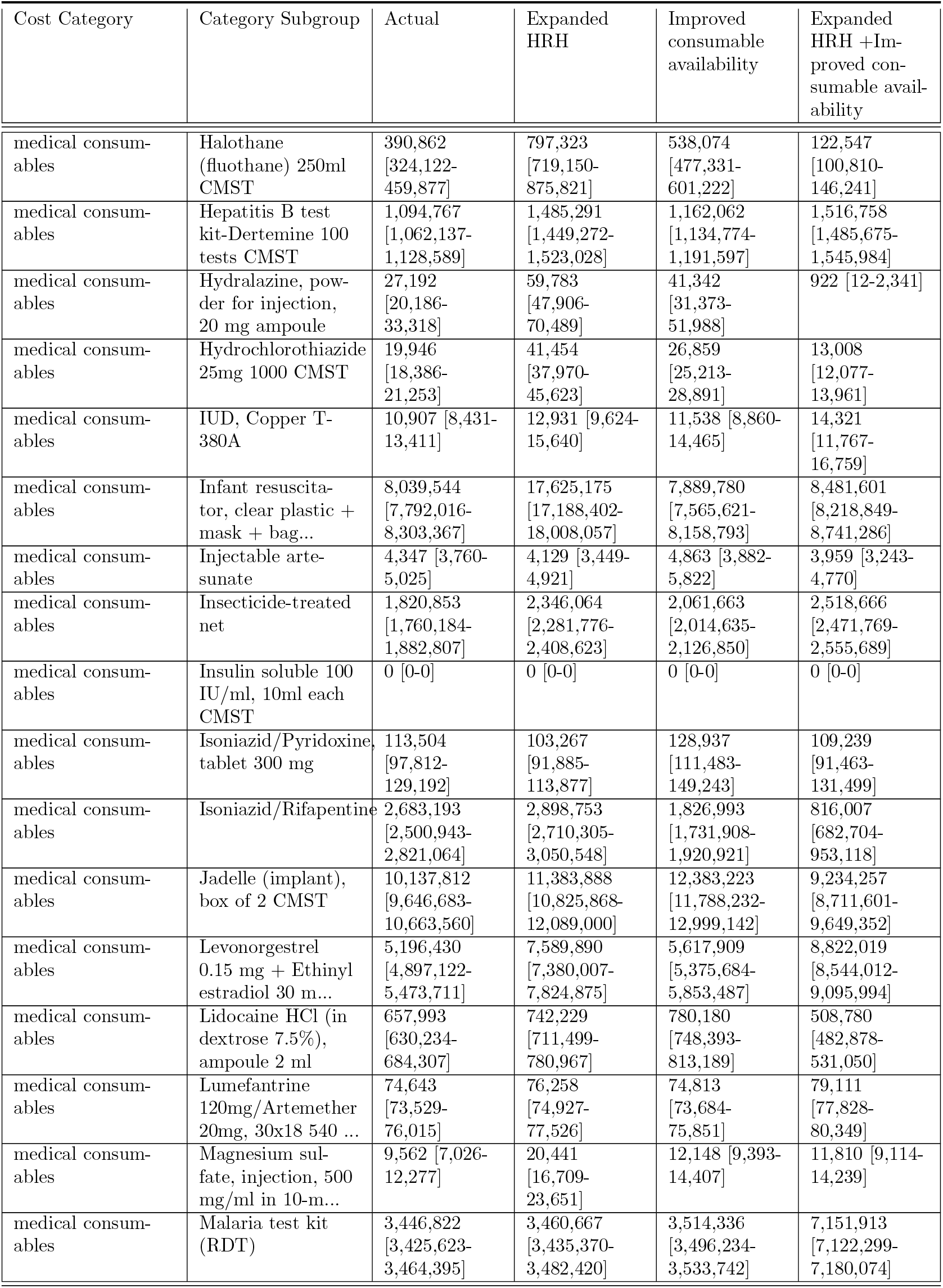
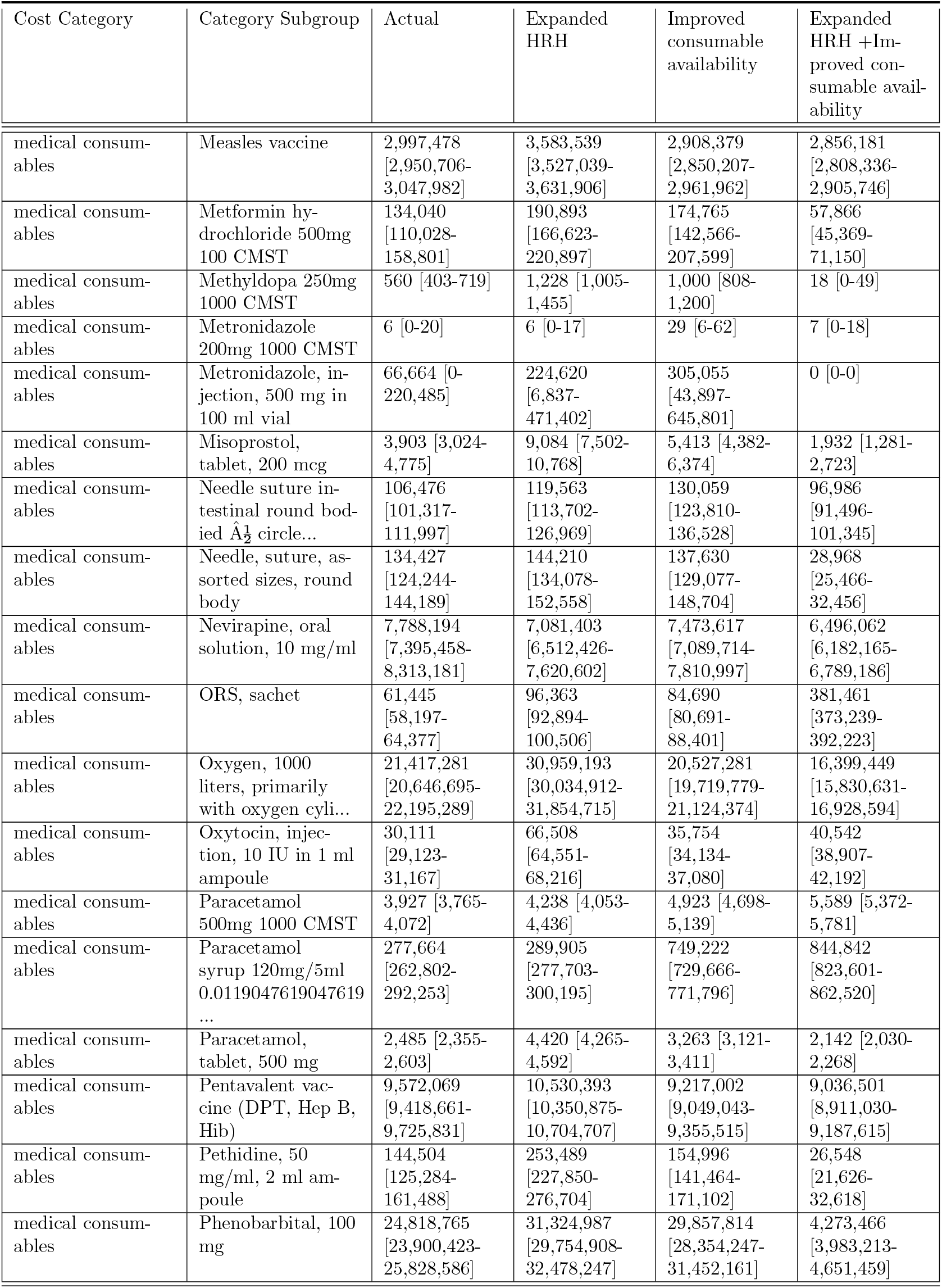
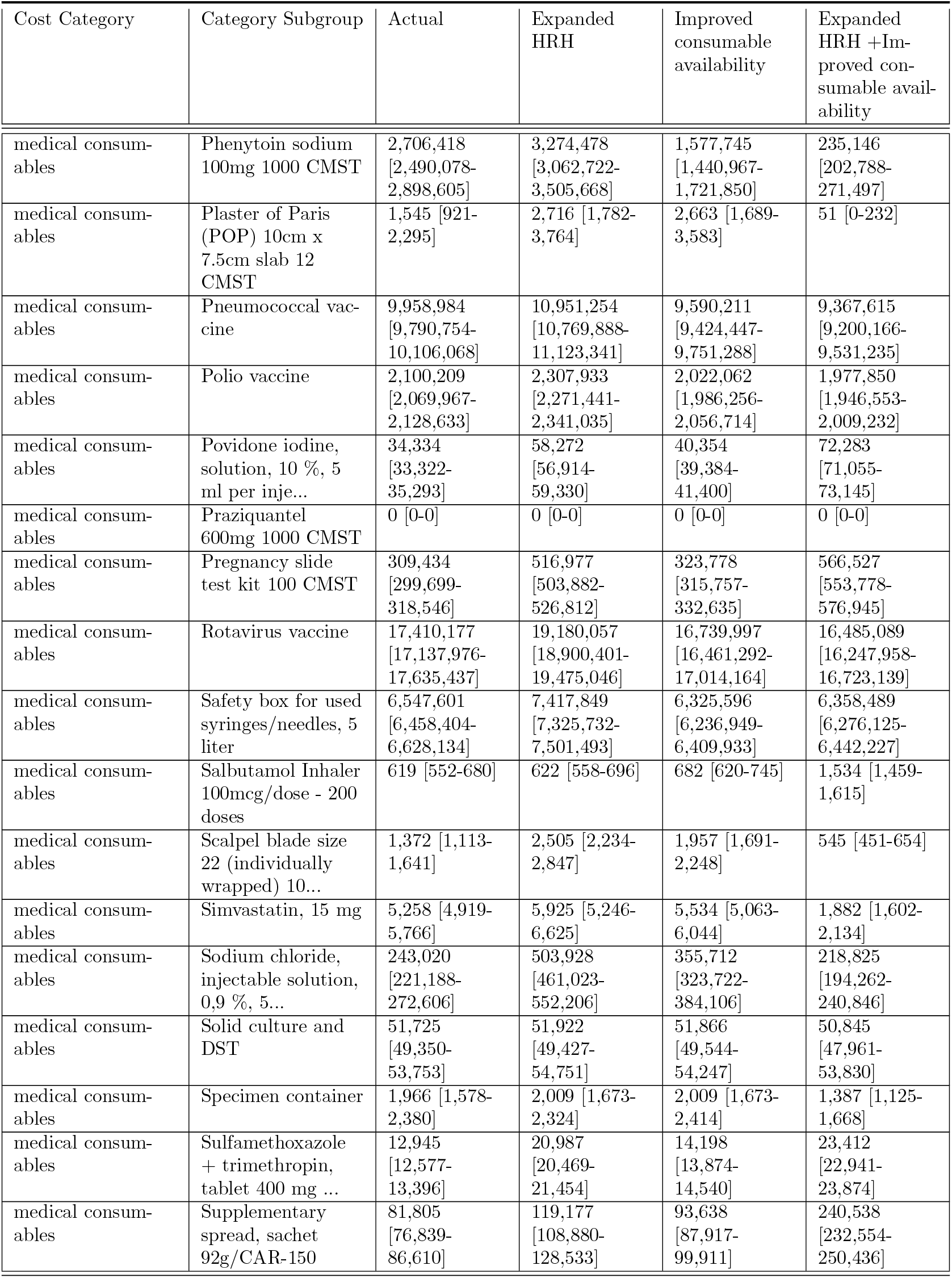
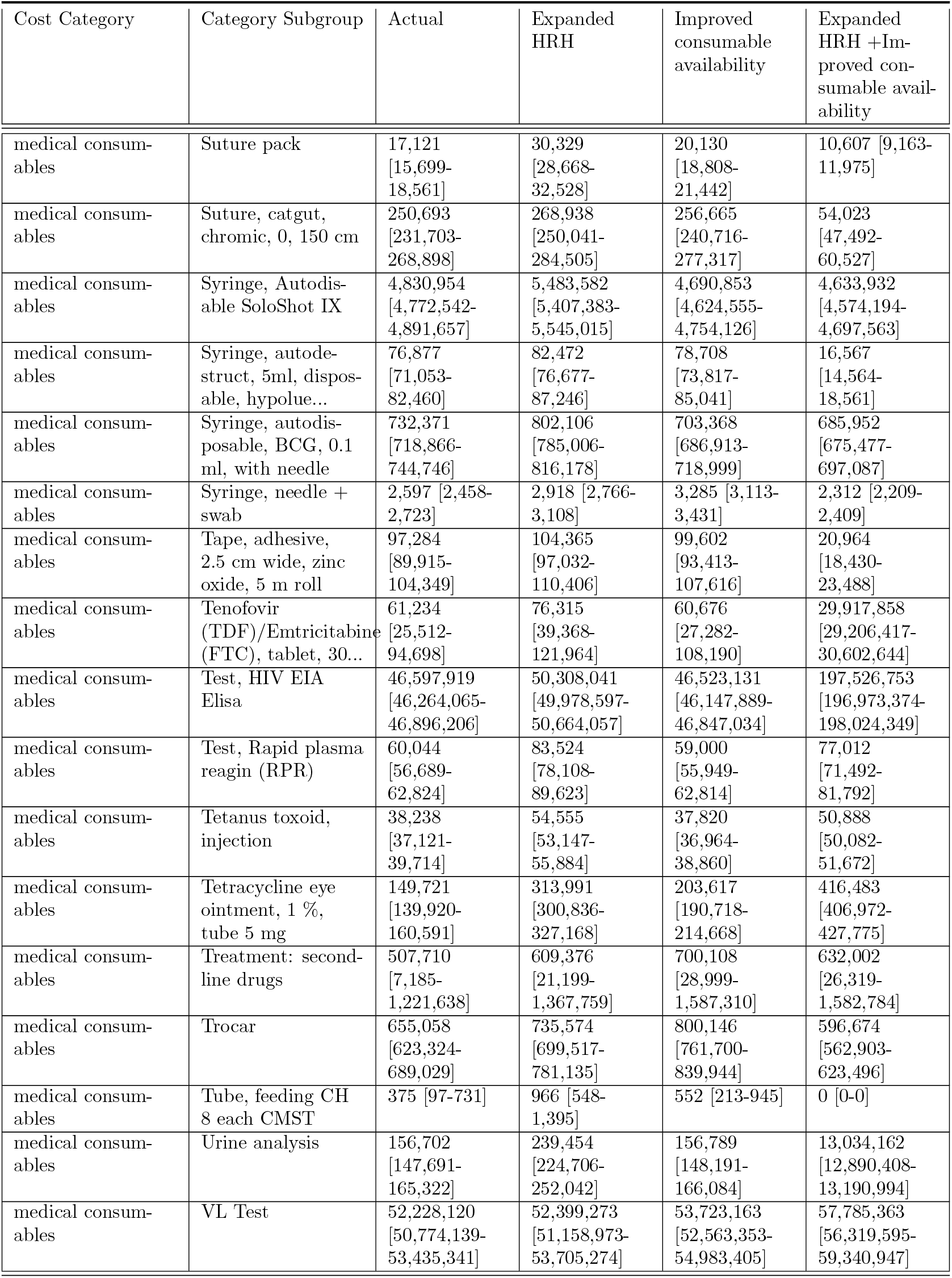
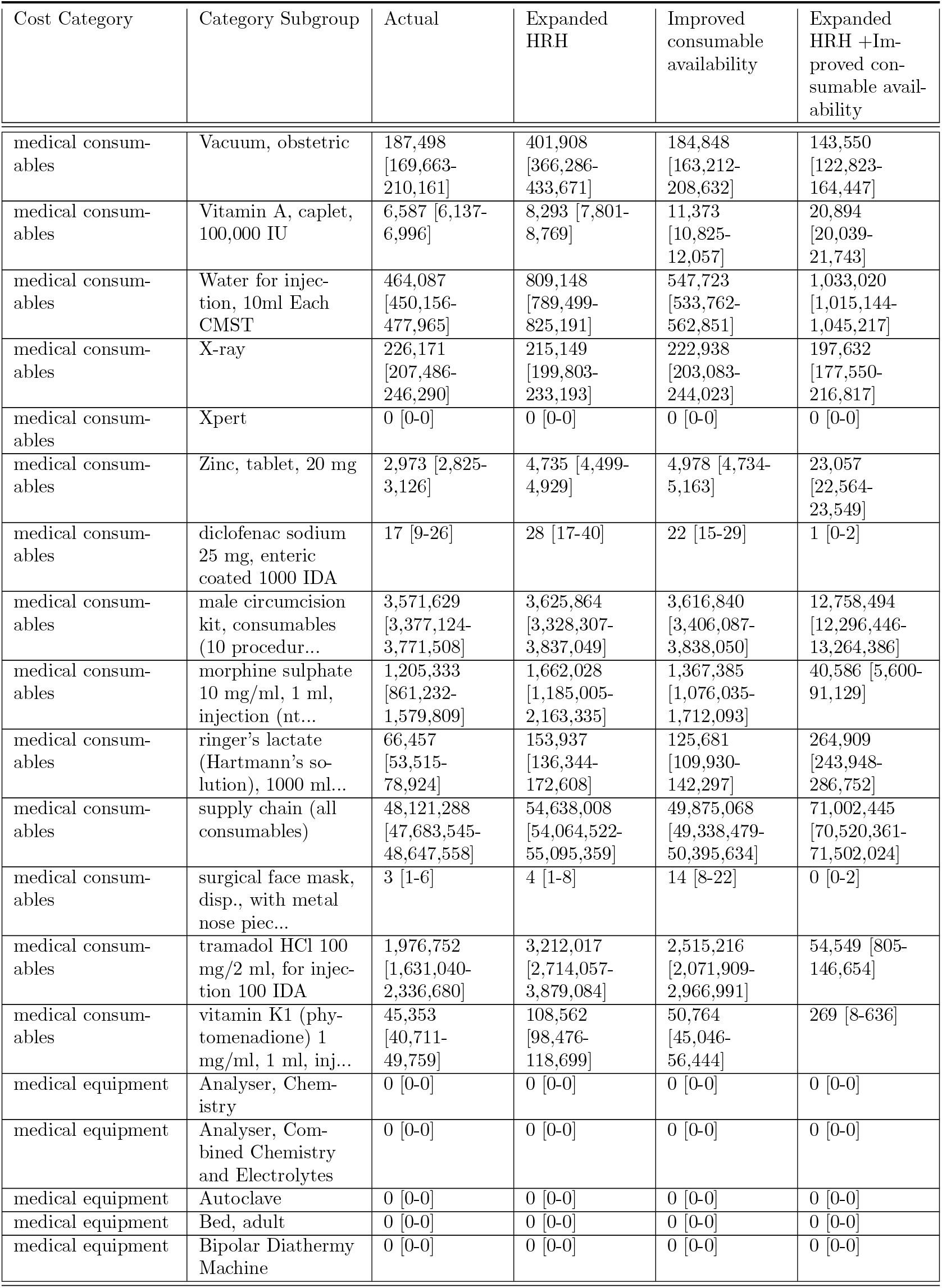
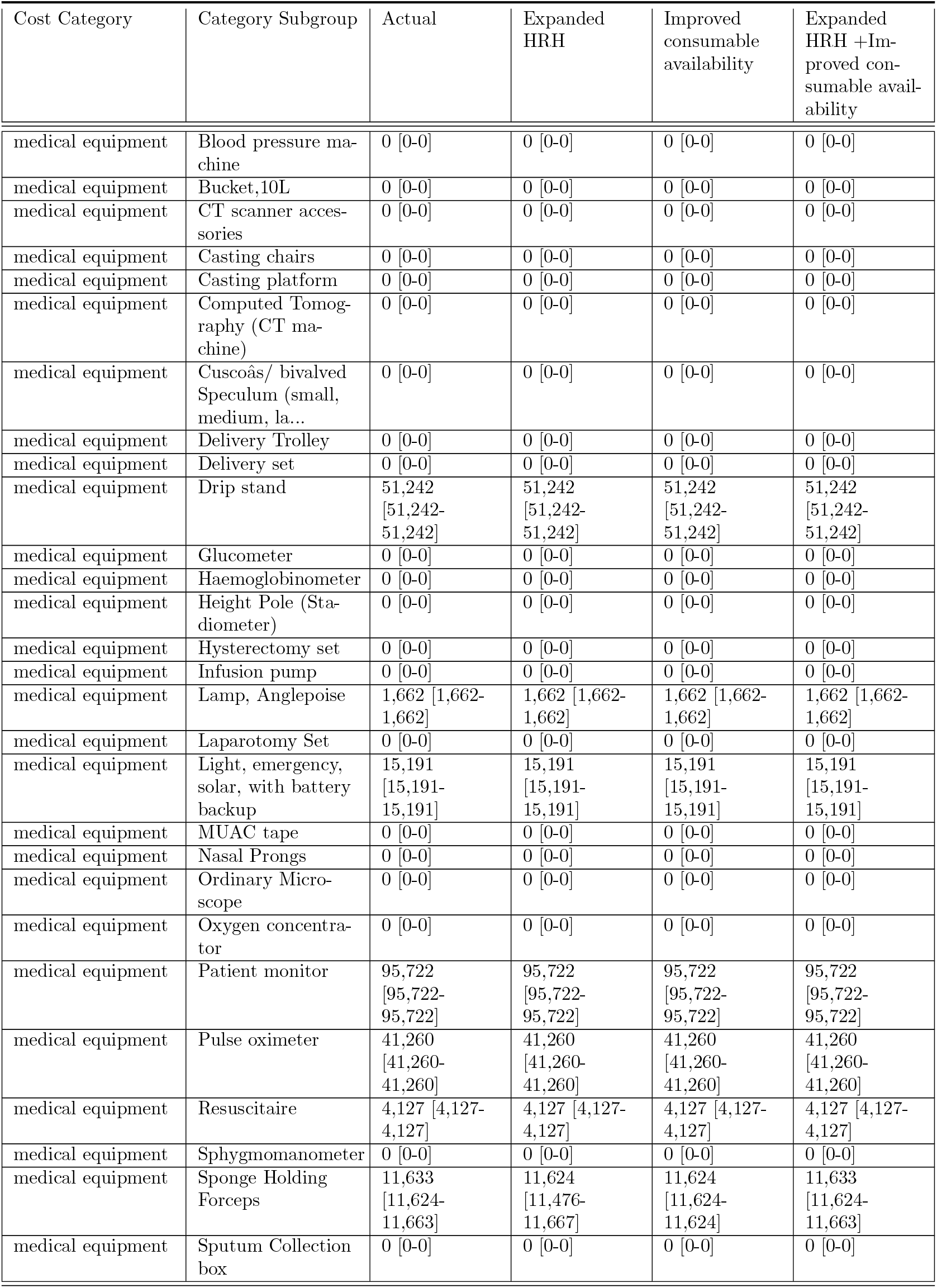
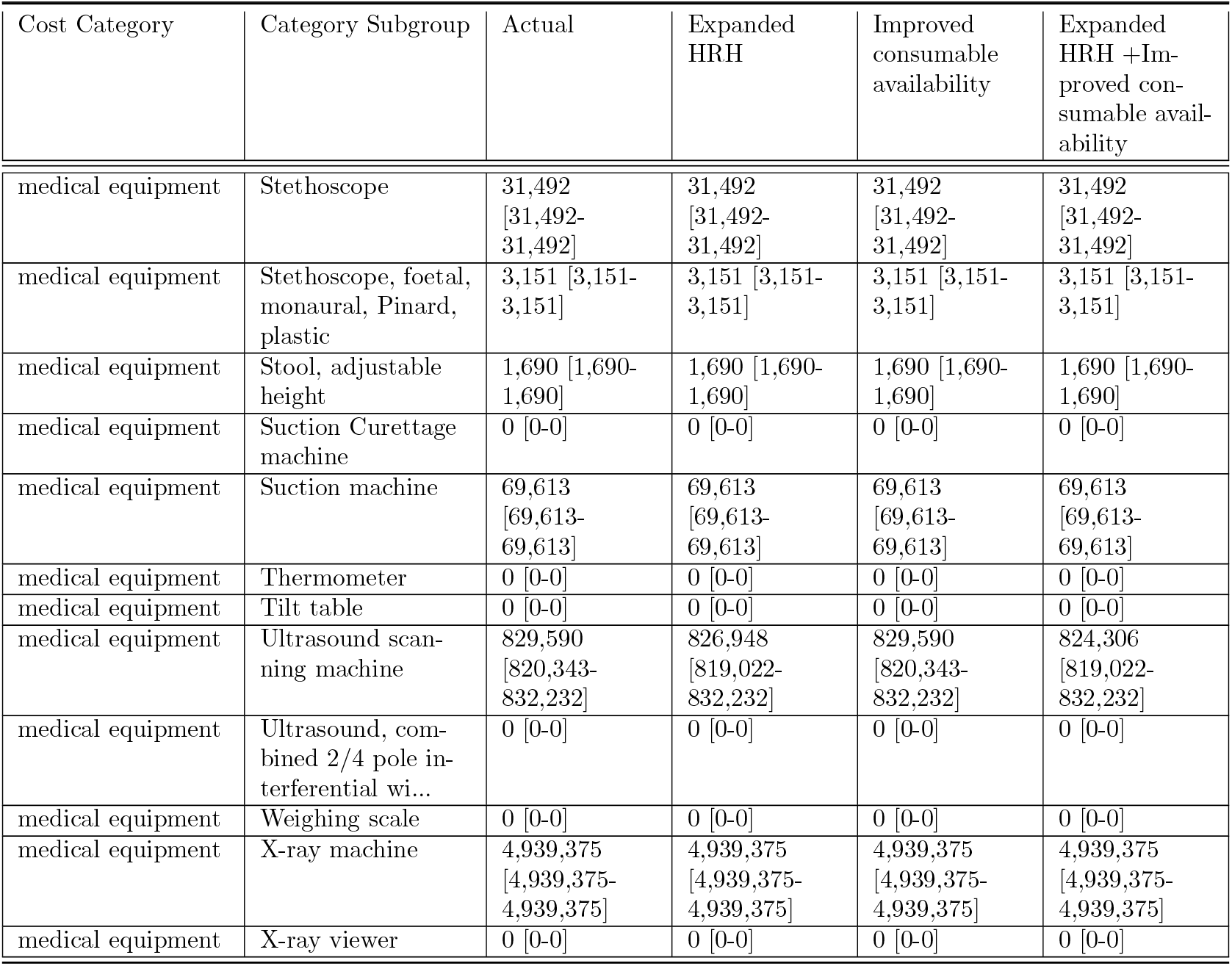
Summarized Costs by Category and Category Subgroup.

#### F.2 Cost by subcategory

**Table F.2:** Summarized Costs by Category and Category Subgroup.

| Cost Category | Cost Subcategory | Actual | Expanded HRH | Improved consumable availability | Expanded HRH +Improved consumable availability |
| --- | --- | --- | --- | --- | --- |
| facility operating cost | building maintenance | 25,294,276<br>[25,294,276-25,294,276] | 25,294,276<br>[25,294,276-25,294,276] | 25,294,276<br>[25,294,276-25,294,276] | 25,294,276<br>[25,294,276-25,294,276] |
| facility operating cost | food for inpatient care | 11,969,939<br>[11,969,939-11,969,939] | 11,969,939<br>[11,969,939-11,969,939] | 11,969,939<br>[11,969,939-11,969,939] | 11,969,939<br>[11,969,939-11,969,939] |
| facility operating cost | fuel for ambulance | 30,439,935<br>[30,439,935-30,439,935] | 30,439,935<br>[30,439,935-30,439,935] | 30,439,935<br>[30,439,935-30,439,935] | 30,439,935<br>[30,439,935-30,439,935] |
| facility operating cost | utilities and maintenance | 89,906,685<br>[89,906,685-89,906,685] | 89,906,685<br>[89,906,685-89,906,685] | 89,906,685<br>[89,906,685-89,906,685] | 89,906,685<br>[89,906,685-89,906,685] |
| facility operating cost | vehicle maintenance | 24,268,997<br>[24,268,997-24,268,997] | 24,268,997<br>[24,268,997-24,268,997] | 24,268,997<br>[24,268,997-24,268,997] | 24,268,997<br>[24,268,997-24,268,997] |
| human resources for health | inservice training cost for cadres used | 57,915,137<br>[57,915,137-57,915,137] | 92,296,067<br>[92,296,067-92,296,067] | 57,915,137<br>[57,915,137-57,915,137] | 106,388,748<br>[106,388,748-106,388,748] |
| human resources for health | mentorship and supportive cost for cadres used | 249,464,590<br>[249,464,590-249,464,590] | 397,557,558<br>[397,557,558-397,557,558] | 249,464,590<br>[249,464,590-249,464,590] | 458,260,600<br>[458,260,600-458,260,600] |
| human resources for health | preservice training and recruitment cost for attrited workers | 13,502,061<br>[13,502,061-13,502,061] | 21,517,468<br>[21,517,468-21,517,468] | 13,502,061<br>[13,502,061-13,502,061] | 24,749,120<br>[24,749,120-24,749,120] |
| human resources for health | salary for cadres used | 513,803,145<br>[513,803,145-513,803,145] | 818,818,911<br>[818,818,911-818,818,911] | 513,803,145<br>[513,803,145-513,803,145] | 943,844,325<br>[943,844,325-943,844,325] |
| medical consumables | cost of consumables dispensed | 387,560,775<br>[373,694,687-401,898,840] | 439,180,994<br>[422,890,566-454,614,731] | 401,893,411<br>[387,199,119-416,936,857] | 570,519,037<br>[557,898,662-583,101,336] |
| medical consumables | cost of excess consumables stocked | 41,125,586<br>[39,339,693-43,002,890] | 47,559,274<br>[45,472,772-49,678,455] | 42,416,421<br>[40,517,407-44,444,335] | 62,003,097<br>[60,421,585-63,649,610] |
| medical consumables | cost of excess separately managed medical supplies | 3,224,639<br>[2,994,729-3,420,553] | 5,064,834<br>[4,747,301-5,381,367] | 3,062,178<br>[2,828,539-3,264,996] | 2,061,055<br>[1,982,035-2,144,709] |
| medical consumables | cost of separately managed medical supplies dispensed | 22,530,744<br>[20,924,350-23,899,608] | 35,388,295<br>[33,169,671-37,599,933] | 21,395,623<br>[19,763,167-22,812,724] | 14,400,713<br>[13,848,596-14,985,211] |
| medical consumables | supply chain | 48,121,288<br>[47,683,545-48,647,558] | 54,638,008<br>[54,064,522-55,095,359] | 49,875,068<br>[49,338,479-50,395,634] | 71,002,445<br>[70,520,361-71,502,024] |
| medical equipment | major corrective maintenance cost annual total | 144,555<br>[144,383-144,606] | 144,506<br>[144,349-144,607] | 144,555<br>[144,383-144,604] | 144,457<br>[144,358-144,606] |
| medical equipment | replacement cost annual total | 2,586,436<br>[2,581,662-2,587,826] | 2,585,067<br>[2,580,841-2,587,830] | 2,586,428<br>[2,581,662-2,587,790] | 2,583,713<br>[2,580,981-2,587,826] |
| medical equipment | service fee annual total | 2,691,806<br>[2,688,360-2,692,791] | 2,690,821<br>[2,687,867-2,692,791] | 2,691,806<br>[2,688,360-2,692,791] | 2,689,837<br>[2,687,867-2,692,791] |
| medical equipment | spare parts annual total | 672,952<br>[672,090-673,198] | 672,705<br>[671,967-673,198] | 672,952<br>[672,090-673,198] | 672,459<br>[671,967-673,198] |

**Figure F.1:**
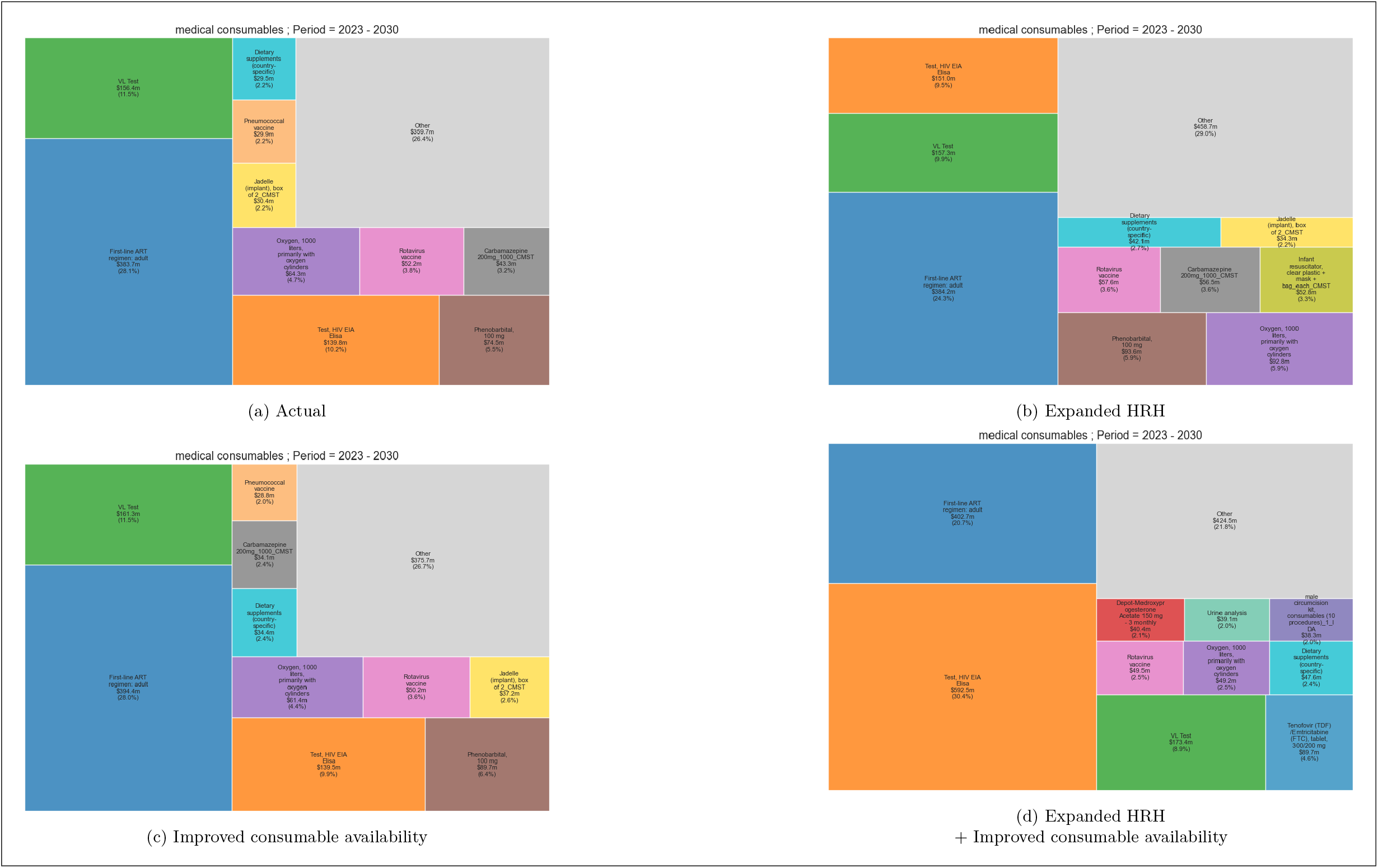
Summary by category subgroup (Medical consumables), 2023-2030. This figure reports the cost over 17 years for the top 10 consumables.

**Figure F.2:**
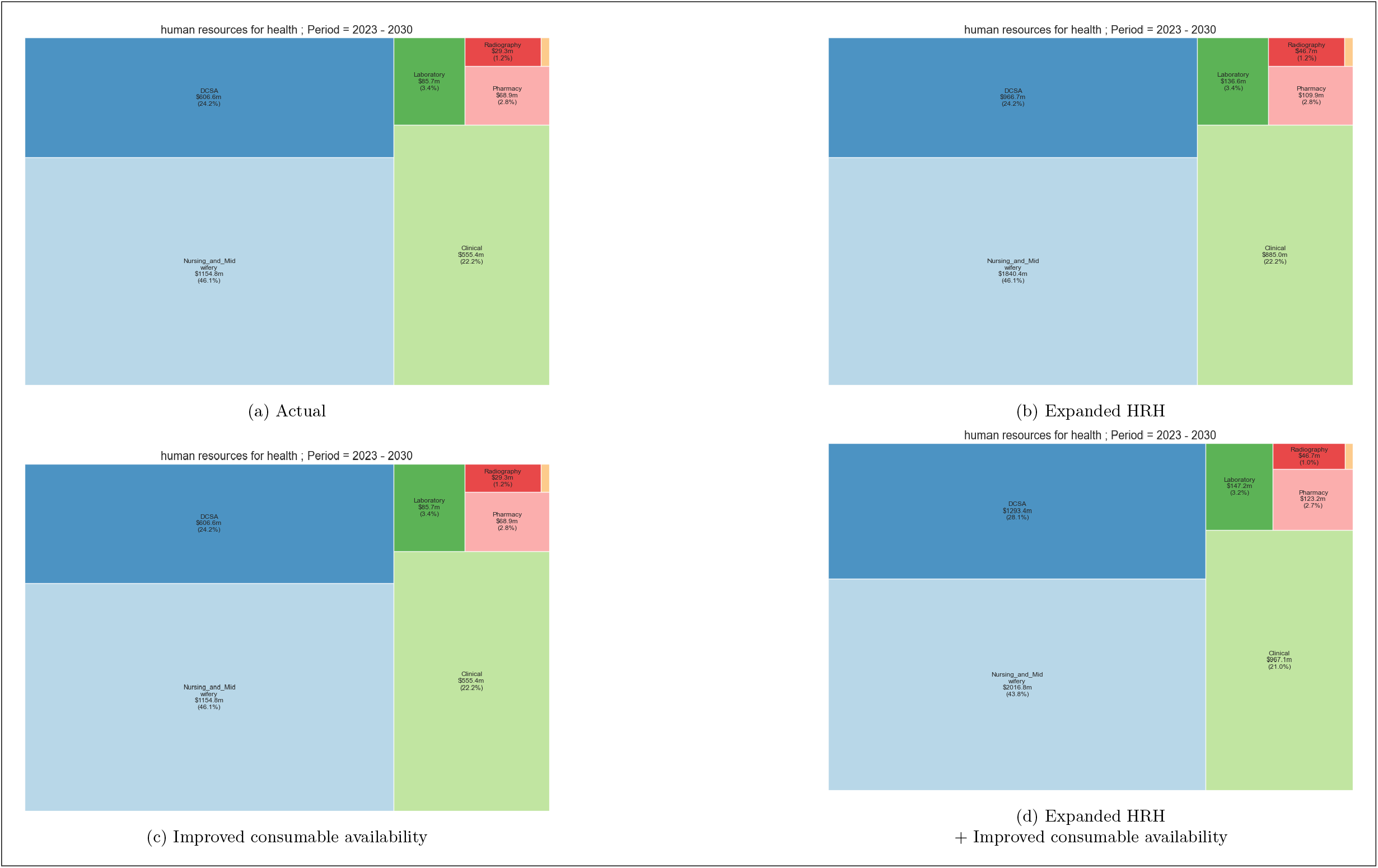
Summary by category subgroup (Human resources for health), 2023-2030. This figure reports the cost over 17 years by health worker cadre.

**Figure F.3:**
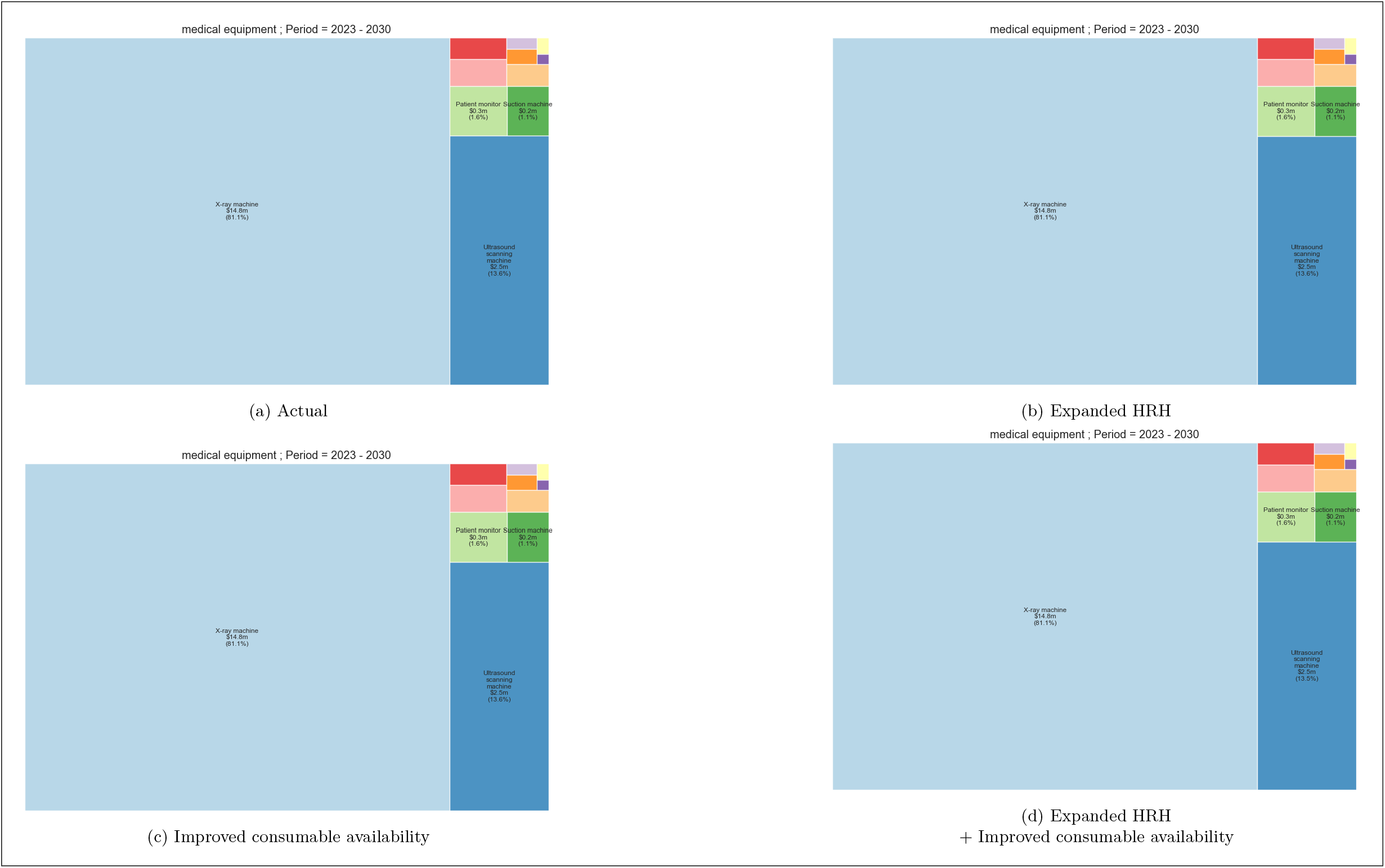
Summary by category subgroup (Medical equipment), 2023-2030. This figure reports the cost over 17 years for the top 10 medical equipment.

**Figure F.4:**
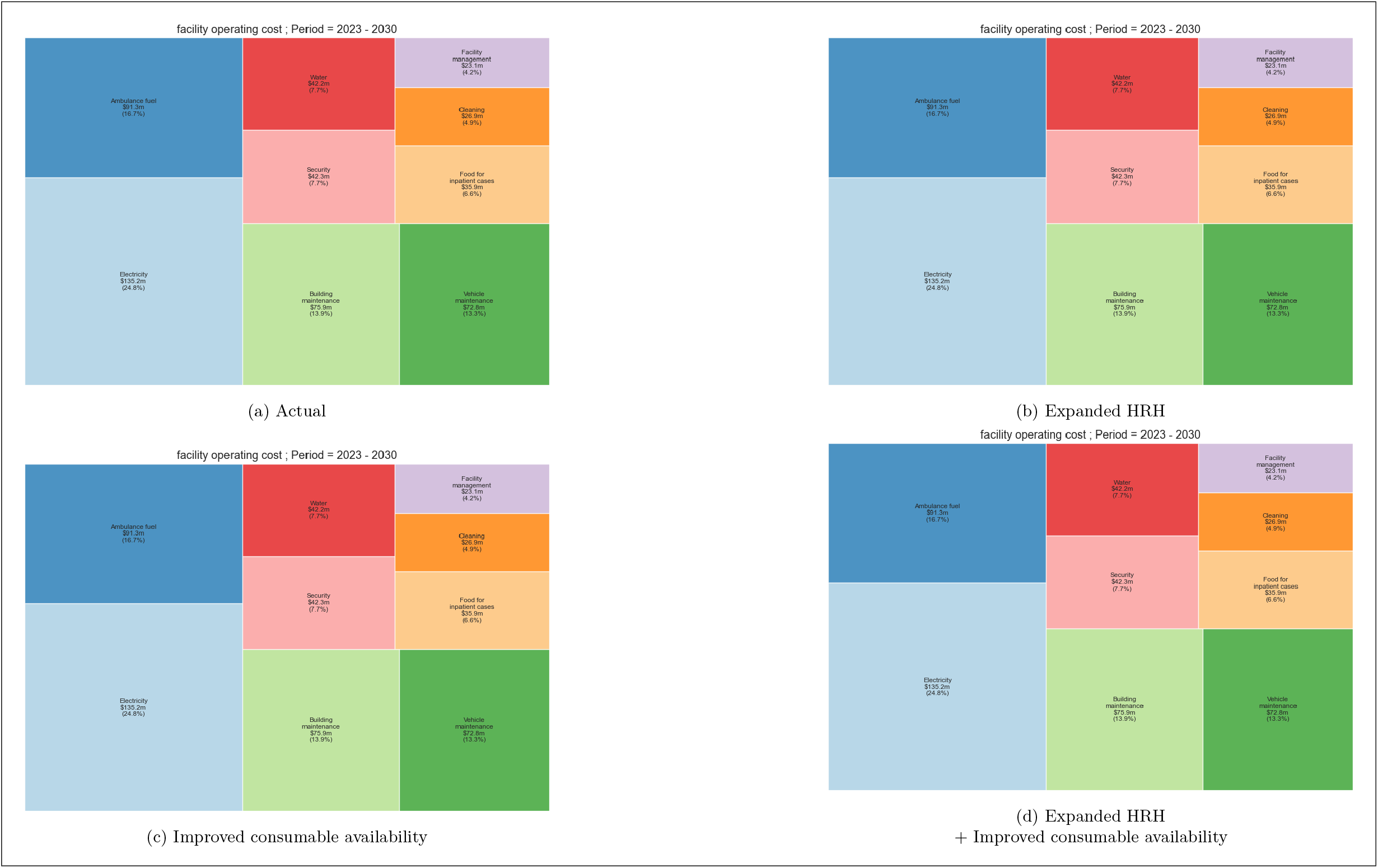
Summary by category subgroup (Facility operations), 2023-2030. This figure reports the cost over 8 years by the type of operating cost.

## Footnotes

1 Although included in the formula, we assume that the proportion of staff hired from abroad in 0.

2 This is different from the more detailed calibration process followed for the epidemiological aspects of the model, details of which can be found in Hallett et al (2024)[5]

3 More recent expenditure information in the granular format provided by the Resource Mapping database is not currently available.

4 For example, the use of Gentamicin sulphate was included for acute lower respiratory illnesses, management of sepsis in mothers and newborns and for management of obstructed labour. Not all conditions which might warrant the use of this antibiotic were modelled.

5 If the actual availability is higher, then no change is made.

## Notes

### Competing Interest Statement

The authors have declared no competing interest.

### Funding Statement

This project is funded by The Wellcome Trust (223120/Z/21/Z). The initial development of the model was completed with support by the UK Research and Innovation as part of the Global Challenges Research Fund (MR/P028004/1). TBH, TM, MM and BS acknowledge funding from the MRC Centre for Global Infectious Disease Analysis (reference MR/X020258/1), funded by the UK Medical Research Council (MRC). This UK funded award is carried out in the frame of the Global Health EDCTP3 Joint Undertaking.

